# Genetic architecture of treatment-resistant schizophrenia across East Asian and European cohorts: insights from GWAS, TWAS, and synaptic pathway analyses

**DOI:** 10.64898/2026.08.06.26359853

**Authors:** Perry BM Leung, Kenneth CY Wong, Sophie Smart, Nick R Zhang, Zoe ZY Zheng, Jinghong Qiu, Edoardo Spinazola, Antonio Pardiñas, Justin D. Tubbs, Amy CY Liu, Karen KY Ho, Koi-Man Cheng, Karen SY Hung, Eric FC Cheung, Vicki HM Ling, Tomy CK Hui, Ole A Andreassen, Thomas R.E. Barnes, Philippe Conus, Benedicto Crespo Facorro, Gillian A Doody, Kim Q. Do, Chin B Eap, Eileen Joyce, Ingrid Melle, Paulo Menez, Craig Morgan, Francis A O’Neill, Baptiste Pignon, Filip Španiel, Ilaria Tarricone, Andrea Tortelli, Alp Üçok, Homero Vallada, Javier Vázquez-Bourgon, The STRATA Consortium, Luis Alameda, Evangelos Vassos, James T.R. Walters, James MacCabe, Marta Di Forti, Robin M Murray, Hon-Cheong So, Pak C Sham, Simon SY Lui

## Abstract

In about a quarter of people with schizophrenia-spectrum disorder (SSD), the illness is unresponsive to standard antipsychotic treatment, yet the biological mechanisms underlying this remain poorly understood. Although such treatment-resistant schizophrenia (TRS) shares a substantial genetic liability with treatment-responsive schizophrenia, the limited efficacy of dopamine antagonists in TRS indicates that mechanisms beyond dopamine signalling likely contribute to treatment-resistance, requiring the identification of alternative biological pathways. This is the first cross-ancestry genetic study to investigate the genetic architecture of TRS, by directly comparing patients with treatment-resistant and treatment-responsive schizophrenia in two independent Hong Kong (N=798) and STRATA-G consortium (N=1, 243) cohorts. Using an integrated multi-level analytic framework, we conducted a genome-wide association study (GWAS) with gene-based and gene set-based analyses, pathway polygenic-risk-scores, and transcriptome-wide association study (TWAS). We further conducted gene-set enrichment analysis focusing on expert-curated synaptic pathways and brain tissues. Genetic signals at the gene, pathway, and genetically predicted expression levels were identified within each ancestry. Whereas limited power constrained individual loci discovery and cross-ancestry concordance, enrichment analyses indicated heterogeneous signals across cohorts, including differences in effect direction, but highlighted cohort-specific, synapse-related biology, particularly pathways involved in presynaptic vesicle dynamics, neurotransmission, and synaptic organization. Collectively, these findings highlight synaptic biology as one potential pathway-level signal from common-variant genetic effects associated with treatment resistance in SSD, despite minimal SNP-level discovery. Our work suggests there is promise in pathway-level and multi-omics approaches to elucidate biologically meaningful heterogeneity within SSD and provides support for synaptic mechanisms as potential targets for understanding and stratifying treatment-resistance.

## 1. Introduction

Schizophrenia-spectrum disorder (SSD) represents a heterogeneous group of psychosis syndromes, with schizophrenia as the prototype (GBD 2019 Mental Disorders Collaborators, 2022) characterised by positive, negative, and cognitive symptoms. SSD is associated with substantial functional impairments and reduced life expectancy (GBD 2019 Mental Disorders Collaborators, 2022; Owen et al., 2016). Dopaminergic and synaptic dysfunction are central to the current biological models for its pathophysiology, with the synaptic hypothesis proposing that aberrant synaptic pruning and plasticity may contribute to altered neural connectivity and downstream striatal dopaminergic hyperactivity (Howes & Onwordi, 2023). While dopamine D2 receptor antagonists effectively reduce positive symptoms and lead to recovery in many SSD patients, approximately a quarter of first-episode SSD patients will develop treatment-resistant schizophrenia (TRS), manifesting persistent symptoms and significant functional impairments despite trials of at least two non-clozapine antipsychotics with sufficient dosage and duration (Siskind et al., 2022). Identifying treatment mechanisms beyond simple D_2_ blockade is needed in this complex group of SSD.

Aberrant synaptic signalling underlying non-dopaminergic mechanisms is increasingly implicated in TRS. Neuroimaging and post-mortem data highlight dysregulated glutamatergic signaling and NMDA receptor dysfunction on GABAergic interneurons (Murray et al., 2026; Potkin et al., 2020). Cerebrospinal fluid proteomics further reveals upregulated immune markers alongside downregulated synaptic and glutamatergic processes (Fischer et al., 2025), while oxidative stress and redox dysregulation have also been linked to these alterations in TRS (Camporesi et al., 2024). Additionally, systemic inflammation and neuroinflammation mechanisms have been proposed as contributors to TRS (Leboyer et al., 2021). Collectively, these findings suggest that TRS extends beyond the dopaminergic model, with downstream alterations in neural circuitry, calling for the investigation of synaptic pathways in TRS.

Clinically, TRS can be broadly classified into early- and late-emerging forms with distinct hypothesised neurobiological mechanisms. Early-emerging TRS has been associated with a distinct dopaminergic profile compared with treatment-responsive schizophrenia, including reduced presynaptic striatal hyperdopaminergia which renders D2-receptor-blocking agents ineffective (Demjaha et al., 2017; Demjaha et al., 2012; Lally et al., 2016). By contrast, late-emerging TRS may reflect progressive neurobiological changes across the illness-course, possibly resulting from multiple prolonged episodes or antipsychotic-induced dopamine supersensitivity (Murray et al., 2026; Potkin et al., 2020). Despite these distinct neurobiological trajectories, clozapine remains the only FDA-approved pharmacological agent for treating TRS (Fenton & Kang, 2023). However, its adverse side effects (e.g., agranulocytosis, myocarditis, pneumonia, gastrointestinal hypomotility, and sedation) (Partanen et al., 2024; Taylor et al., 2025), and strict blood monitoring requirements limit its use and contribute to prescribing hesitancy (Whiskey et al., 2021). This clinical reality highlights the need to better understand the neurobiological mechanisms of TRS for supporting the development of safer and more effective treatments.

Advances in psychiatric genetics have provided an important avenue to dissect treatment-response heterogeneity. The largest genome-wide association study (GWAS) meta-analysis of TRS to-date, using genome-wide SNP-by-treatment-response interaction models, revealed a near-perfect genetic correlation (r ≈ 0.97) between TRS and treatment-responsive SSD, suggesting substantial shared common-variant polygenic risk (Pardiñas et al., 2022). Although SNP-based interaction models identified modest heritability (∼1–4%) and negative correlations with cognitive performance, a well-established clinical predictor of poor antipsychotic response (Millgate et al., 2023), no single locus reached genome-wide significance (Pardiñas et al., 2022). This lack of locus-level discovery may stem from limited effective sample size, high polygenicity, phenotype misclassification, and cohort heterogeneity, or may reflect genuine modest common-variant heritability, highlighting the challenge of SNP-level discovery and motivating downstream approaches that aggregate signals across genes, pathways and transcriptomic regulation.

Gene- and pathway-based approaches aggregate small variant effects into interpretable biological processes (de Leeuw et al., 2015), and prior schizophrenia GWAS have consistently shown enrichment of synaptic signaling pathways (Schijven et al., 2018). Furthermore, because linkage disequilibrium and allele frequencies vary across populations, individual SNP associations often fail to transfer across diverse ancestries (Martin et al., 2019). To address this, pathway-level aggregation captures conserved biology across populations better than single-SNP analyses (Smith et al., 2022; Zhang et al., 2023). In this study, we addressed genetic heterogeneity in TRS through two complementary aims across two independent multi-ancestry cohorts. First, we evaluated the similarity of common-variant signals associated with TRS in two independent cohorts of different ancestry and ascertainment, across single nucleotide polymorphism (SNP), gene, polygenic risk score (PRS) and genetically predicted gene-expression biological signals. Second, we tested whether TRS-associated genetic liability was preferentially enriched in synaptic pathways, based on the hypothesis that TRS may involve synaptic and circuit-level disruption beyond dopaminergic dysfunction (Murray et al., 2026).

## 2. Method

### 2.1 The East Asian sample gathered from Hong Kong

We recruited a total of 803 patients with SSD from psychiatric services in Hong Kong. This sample comprised two non-overlapping cohorts: (i) 620 SSD patients from the well-established early psychosis intervention service at Castle Peak Hospital in Hong Kong (HK) between 2009 and 2021 (Wong et al., 2024), comprising a mix of TRS and non-TRS cases at the time of this study and (ii) 183 patients with TRS receiving clozapine from adult psychiatric clinics at Castle Peak Hospital and Queen Mary Hospital between 2022 and 2023, representing a clinically distinct group with chronic illness and confirmed treatment-resistance. Inclusion criteria for both cohorts were: (1) age ≥18 years, (2) Chinese ethnicity, (3) a clinical diagnosis of schizophrenia or schizoaffective disorder, and (4) provision of a blood sample for DNA sequencing. Patients with incomplete medication records or lost to follow-up before 2022 were excluded due to unavailable electronic medical records. Clinical diagnoses were based on ICD-10 criteria recorded in electronic medical records and were confirmed using DSM-IV criteria through structured interviews conducted by qualified psychiatrists during routine care. TRS cases were first identified based on a history of clozapine prescription, and further confirmed by documented non-response to at least two different antipsychotics administered at chlorpromazine-equivalent doses of ≥600 mg/day for a minimum of 4–6 weeks each (Howes et al., 2017). We also extracted clinical information, including age-at-onset and medication history. Given the differences in recruitment setting and clinical profile, cohort-specific characteristics are presented in **Supplementary Table 1**, and potential batch/cohort effects were rigorously adjusted for in all downstream genetic analyses. This study was approved by the institutional review boards of the Hong Kong Hospital Authority and participating hospitals (CPH: NTWC/CREC/823/10, NTWC/CREC/1293/14, CIRB-2023-025-4; QMH: UW-22-724), and written informed consent was obtained from all participants.

### 2.2 The European sample gathered from the ‘Schizophrenia: Treatment Resistance and Therapeutic Advances’ (STRATA) Consortium

The STRATA-Genetics (STRATA-G) study combined individual-level genetic and clinical data from first-episode psychosis (FEP) patients across 11 European and Brazilian cohorts between 1996 and 2014 (Pardiñas et al., 2022), prior to the development of the TRRIP guidelines for TRS. While several cohorts restricted inclusion to SSD, others encompassed transdiagnostic FEP presentations, including non-schizophrenia psychoses. Participants were followed prospectively for at least 12 months. Detailed descriptions of the design and sample characteristics of each contributing cohort have been reported elsewhere (Smart et al., 2022). The consortium defined TRS patients based on the following criteria: (1) clozapine prescription; or (2) non-response to two different antipsychotics trials with dosages in at least the mid-point of the licensed therapeutic range for at least 6 weeks; or (3) persistent psychotic symptoms and moderate functional impairment despite two different antipsychotics trials (Smart et al., 2022).

### 2.3 Quality control (QC) procedures

The DNA extractions and sequencing details for the Hong Kong cohort are provided in Supplementary Materials. In brief, genomic DNA was extracted from peripheral blood and sequenced using low-coverage whole-genome sequencing (∼10×mean depth). Variant calling and QC followed GATK pipeline, including variant quality score recalibration (VQSR) and genotype refinement. Pre-imputation filtering retained high-quality sites with variant quality score ≥30, alternative allele depth ratio ≥0.2, minor allele frequency (MAF) ≥1%, and Hardy-Weinberg equilibrium (HWE) p≥1x10⁻⁴. The alternative allele depth ratio filter was applied to exclude variants with highly imbalanced read support, which are more likely to reflect sequencing artefacts. A relaxed HWE threshold was used to preserve haplotype structure. Genotypes were subsequently phased using ShapeIt4 and imputed with Beagle5 with the East Asian subset of the 1000 Genomes reference panel. Post-imputation variants were retained when imputation quality R²>0.8. Genotypes were converted to hard calls using a posterior probability threshold of 0.9 to ensure high-confidence genotype assignments for downstream analyses. GWAS QC retained SNPs with missingness <0.05 (i.e. geno 0.05), MAF >1%, and HWE p>1x10⁻⁶, followed by LD pruning (window=50, step=5, r²<0.2) (Anderson et al., 2010). The first four within-sample principal components were selected based on the elbow plots and reported in PCA scatter plots (See **Supplementary Figures 15-17**) to control batch effects and population structure, and no ancestry substructure was observed in our HK sample. The final post-QC sample comprised 798 SSD cases (290 TRS, 508 non-TRS). Detailed QC procedures are summarised in **Supplementary Figure 2**.

The QC procedures for each STRATA-G cohort have been described elsewhere (Pardiñas et al., 2022). Briefly, included participants were genotyped on Illumina platforms, allowing up to 5% of missingness at both marker and individual levels. Imputed variants using Michigan Imputation Server were retained with genotype probability >90%, imputation R²>0.8, MAF >1%, and HWE mid-p>1x10⁻⁴. Participants with missing sex, TRS status, or genetic data were excluded. Genetic principal components were recomputed for the STRATA-G comparison sample (N=1,256). After QC, 1,243 individuals remained, including 168 TRS cases.

### 2.4 Statistical analyses

Demographic characteristics were compared between TRS and non-TRS groups in each dataset using chi-square and Wilcoxon rank sum tests as appropriate. Recognising limited sample size constraints, this study integrated GWAS, gene-based, pathway-level, polygenic risk score (PRS), and transcriptome-wide association analyses (TWAS) to aggregate polygenic signals across East-Asian and European cohorts, followed by cross-ancestry meta-analysis. Analysis workflow is summarised in **Figure 1**.

**Figure 1.**
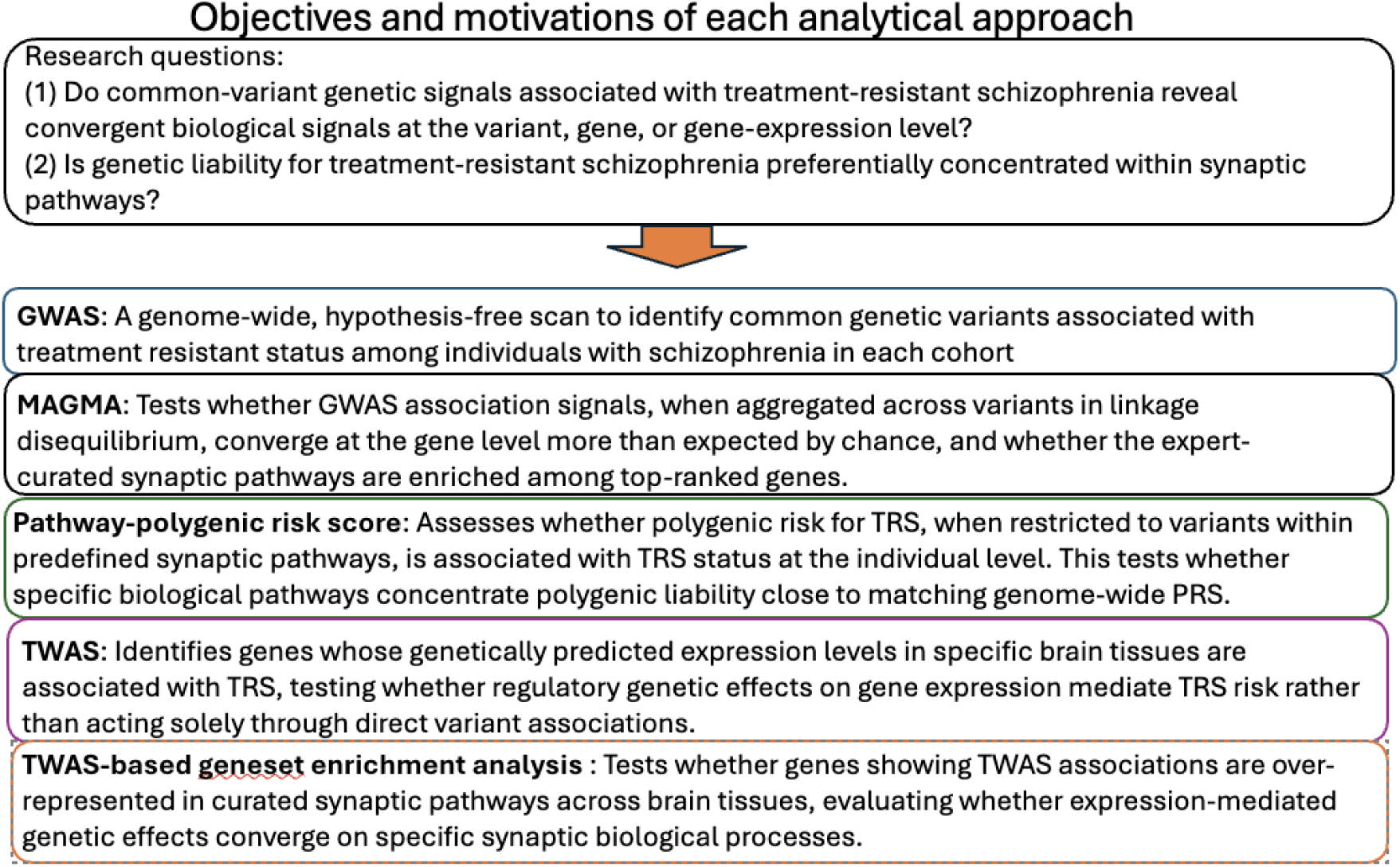
Methods used and their motivations.

#### 2.4.1 GWAS and cross-ancestry meta-analysis

Additive case-case GWAS models compared treatment-resistant and treatment-responsive psychosis cases within each cohort (SSD only in HK; non-schizophrenia psychosis included in STRATA-G), adjusting for sex, batch/cohort, and the first four genetic principal components. Index SNPs were identified by LD clumping using ancestry-matched 1000 Genomes Phase 3 reference panels.

Because individual cohort sample sizes limit power for single-variant detection and cross-cohort genetic correlation (e.g., LD Score Regression), summary statistics were combined using fixed-effects meta-analysis in METAL (Willer et al., 2010). An unfiltered meta-analysed summary dataset retaining all harmonisable variants was generated to maximise genomic coverage, alongside a heterogeneity-filtered HETQC dataset excluding variants with cross-study heterogeneity (HetP < 0.05). Meta-analysis LD clumping used a reference panel combining the European and East Asian populations in 1000 Genomes Phase 3.

#### 2.4.2 Gene set and pathway curation

Synaptic pathways were analysed as hypothesis-driven analysis using SynGo, an expert-curated synaptic ontology comprising 2,922 annotations across 1,112 genes and 259 pathways spanning synaptic compartments and processes (Koopmans et al., 2019). Following standard recommendations, gene-sets were filtered to retain pathways with 15-500 genes to balance biological specificity and statistical stability (Broad Institute, 2019; Fass et al., 2024), yielding 107 retained synaptic pathways (see **Supplementary Table 16**). Pathway overlap was quantified using the Jaccard similarity index, which was computed as the number of intersecting genes divided by the number of union genes between two gene-sets (Fuxman Bass et al., 2013), and visualized as an undirected network graph, with nodes showing overlapping pathways. A Jaccard similarity threshold of ≥25% was selected to highlight substantial gene-set overlap indicative of shared functional themes, while avoiding excessive network density driven by minor or incidental overlaps (see **Supplementary Figure 1**).

In secondary analyses, we evaluated an additional 127 non-synaptic Gene Ontology Biological Process (GO:BP) pathways related to brain cellular structures (indexed by keywords ‘Axon*’, ‘Dendrite’, ‘Glial’, ‘Neuron*’), including multiple highly schizophrenia-relevant pathways on dopaminergic neuron and oxidative stress (see **Supplementary Table 17**). Identical filtering criteria (15–500 genes; Jaccard index) were applied. These primary (SynGo) and secondary (GO:BP) gene sets were applied consistently across all downstream pathway-level analyses introduced below.

#### 2.4.3 Gene-based and gene-set analyses by MAGMA

Gene-based and gene-set analyses were performed using MAGMA (de Leeuw et al., 2015). SNPs were mapped to genes using 35 kb upstream and 10 kb downstream transcription windows (Watson et al., 2019), and gene-level statistics were computed using the SNP-wise multi-marker model with ancestry-matched LD reference panels from 1000 Genomes Phase 3. Competitive gene-set analyses tested enrichment in the curated primary and secondary pathways, accounting for gene size, SNP density, and LD structure. Analyses were performed within each cohort and meta-analysed using MAGMA’s built-in framework, with meta-analysed gene statistics used for competitive gene-set testing. As MAGMA is a competitive enrichment rather than an effect-size model, it does not provide directly comparable effect estimates across cohorts. Therefore, formal cross-cohort heterogeneity testing was not performed.

#### 2.4.4 Polygenic risk score (PRS) calculation

Schizophrenia PRS were calculated using ancestry-matched European (N=320,404) and East Asian (N=58,140) schizophrenia GWAS summary statistics from the PGC as discovery samples (Lam et al., 2019; Trubetskoy et al., 2022). East Asian reference data was applied because matching population ancestry has been shown to improve PRS prediction performance in an East Asian schizophrenia cohort (Lim et al., 2023). TRS-PRS were calculated using summary statistics from the genome-wide SNP-by-treatment-response interaction study by Pardiñas et al. (2022), which was completely independent of our target sample. PRS were generated using PRS-CS and PRS-CSx with ancestry-matched 1000 Genomes LD reference panels following recommended procedures (Ge et al., 2019; Ruan et al., 2022). For the predominantly European STRATA-G sample, PRS-CS was applied using European GWAS summary statistics and a European LD reference panel. For the Hong Kong sample, PRS-CS was applied separately using European and East Asian GWAS summary statistics respectively, while PRS-CSx was used to integrate both ancestry-specific GWAS to obtain PRS_EAS+EUR_-SCZ. All PRS were standardised and tested for association with TRS using logistic regression adjusted for sex, cohort/batch, and principal components.

Pathway-specific PRS (pPRS) were constructed using PRSet (Choi et al., 2023) restricting variants to synaptic gene-sets. Unlike genome-wide PRS, this approach restricts scoring variants to biologically annotated pathways, clarifying whether TRS-associated polygenic burden would be concentrated in specific synaptic processes. PRSet was applied using clumping-and-thresholding with default parameters, and proxy SNPs were included at an LD threshold of r² ≥ 0.8 to allow strongly linked SNPs (Tubbs et al., 2023). Pathway-specific PRS were standardised and analysed using the same regression models as the genome-wide PRS.

#### 2.4.5 Transcriptome-Wide Association Study (TWAS)

TWAS was performed using PrediXcan across 13 GTEx v8 brain tissues. PrediXcan uses genotype-based expression prediction models trained in reference transcriptomic datasets to estimate genetically regulated gene expression and test its association with phenotype (Gamazon et al., 2015). Analyses were restricted to genes with acceptable prediction performance in the reference panel, defined as cross-validated R² > 0.01. Within each cohort and tissue, rank-based inverse normal transformation (INT) was applied to predicted gene expressions to stabilise the distributions of the predicted expression levels and to improve comparability across cohorts (Araujo et al., 2023). Associations with TRS were tested using regression models adjusted for sex, cohort/batch, and principal components. Tissue-specific results were meta-analysed across cohorts using random-effects models to account for potential heterogeneity due to ancestry differences in LD structure, prediction model performance, and cohort-specific estimation noise. Meta-analysis was performed separately for each brain tissue.

#### 2.4.6 Gene set enrichment analysis

Gene-set enrichment analysis was performed using tissue-specific TWAS Z-statistics within each cohort and the meta-analysis. For each brain tissue, genes were ranked by Z-scores, and rank-based competitive GSEA tested whether genes within each pathway were enriched toward the top or bottom of the ranked gene list compared with genes outside the pathway (Subramanian et al., 2005). This yields enrichment scores and p-values for each pathway, indicating whether predicted pathway up- or down-regulation is associated with TRS status.

#### 2.4.7 Additional and sensitivity analysis

To assess whether enrichment signals were preferentially concentrated in synaptic pathways rather than reflecting general brain-wide enrichment, we conducted an additional analysis comparing primary synaptic results against the broader panel of brain-related GO:BP pathways using the same competitive testing framework and multiple-testing corrections.

Two sensitivity analyses were conducted. First, STRATA-G analyses were repeated after restricting the sample to principal component analysis (PCA) defined European ancestry, based on projection onto 1000 Genomes reference populations. Second, given that INT constrains inference to relative and rank-based effects of genetically predicted expression which sacrifices interpretability of absolute effect sizes, TWAS meta-analyses were repeated without INT to evaluate the robustness of association signals when absolute effect sizes were retained.

All analyses were conducted on GRCh38 coordinates. QC and GWAS used PLINK2 (v2.00a3). MAGMA, PRS-CS, PRS-CSx, PRSet, PrediXcan, and METAL follow their own documentation. GSEA was performed using *fgsea* (R v4.1.2). Multiple testing was controlled using FDR (Benjamini & Hochberg, 1995), with FDR<0.05 considered significant and FDR<0.1 considered suggestive.

## 3. Results

The characteristics of the HK and STRATA-G cohorts are summarised in **Supplementary Tables 1 and 2**. The number of TRS participants in each individual cohort is shown in **Supplementary Table 3**. TRS patients had an earlier age-at-onset in both cohorts, and a higher prevalence of self-reported family history of psychosis than non-TRS patients in the HK cohort.

### 3.1 GWAS

Cohort-specific GWAS and cross-cohort meta-analyses were conducted primarily to provide summary statistics for downstream analyses. No variant reached genome-wide significance in the meta-analysis. Six and seven independent index SNPs reached the suggestive significance threshold (p < 1x10⁻⁵) in the HK and STRATA-G cohorts respectively, but showed limited overlap across cohorts, given the modest sample sizes and expected population-specific heterogeneity in LD structure and allele frequencies. Meta-analysis of the two cohorts identified eight independent index SNPs with p < 1x10⁻⁵. As loci are selected based on low p-values in underpowered analyses, effect size estimates at these top hits may be inflated (i.e. winner’s curse). Full GWAS results are provided in **Supplementary Tables 4** and **Supplementary Figures 3-8**.

### 3.2 MAGMA

In MAGMA gene-based analysis, one gene, *OSBPL9*, reached FDR-significance (P_FDR_ < 0.05) in the HK cohort (see **Supplementary Table 5**). In gene-set analysis, *regulation of Synaptic Vesicle Exocytosis* was the only significant pathway (Beta = 0.56, P_FDR_ = 0.007) in gene-set analysis in the STRATA-G cohort. In the meta-analysed data, none of the individual genes or gene-sets reached the suggestive significance threshold of P_FDR_ < 0.1. Detailed results are shown in Table 1 and **Supplementary Table 6**.

**Table 1.** Top-ranked synaptic pathways identified by MAGMA gene set-based analysis in Hong Kong and STRATA-G samples (nominal P < 0.05)

| Geneset | Number of genes | Beta | P | FDR-P |
| --- | --- | --- | --- | --- |
| **(A) MAGMA geneset-based analysis results in Hong Kong sample** |  |  |  |  |
| Neurotransmitter receptor transport | 17 | 0.49 | 0.003 | 0.30 |
| Presynaptic cytosol | 51 | 0.27 | 0.006 | 0.30 |
| Integral component of presynaptic membrane | 123 | 0.16 | 0.01 | 0.44 |
| Trans-synaptic signaling | 207 | 0.11 | 0.03 | 0.45 |
| Postsynaptic endocytic zone | 20 | 0.32 | 0.03 | 0.45 |
| Postsynapse to nucleus signaling pathway | 15 | 0.34 | 0.04 | 0.45 |
| Postsynaptic signaling pathway | 17 | 0.32 | 0.04 | 0.45 |
| Presynaptic membrane | 153 | 0.11 | 0.04 | 0.45 |
| Postsynaptic modulation of chemical synaptic transmission | 34 | 0.23 | 0.05 | 0.45 |
| Postsynaptic process involve in chemical synaptic transmission | 39 | 0.22 | 0.05 | 0.45 |
| **(B) MAGMA geneset-based analysis results in STRATA-G sample** |  |  |  |  |
| **Regulation of synaptic vesicle exocytosis** | **33** | **0.56** | **8x10<sup>-5</sup>** | **0.008** |
| Synaptic vesicle exocytosis | 75 | 0.20 | 0.02 | 0.79 |
| Synaptic vesicle cycle | 202 | 0.10 | 0.03 | 0.79 |
| Process in the presynapse | 288 | 0.09 | 0.03 | 0.79 |
| Regulation of postsynapse assembly | 56 | 0.19 | 0.04 | 0.79 |
| **(C) MAGMA geneset-based analysis results in meta-analysed sample (Hong Kong + STRATA-G)** |  |  |  |  |
| Regulation of synaptic vesicle exocytosis | 35 | 0.33 | 0.009 | 0.72 |
| Process in the presynapse | 299 | 0.08 | 0.04 | 0.72 |
| Presynaptic membrane | 157 | 0.11 | 0.04 | 0.72 |
| Presynaptic cytosol | 51 | 0.18 | 0.05 | 0.72 |

Footnote: No pathways survived FDR correction in the HK or meta-analysed samples

### 3.3 Polygenic-risk-scores (PRS)

Using ancestral-aware PRS-CSx in the HK sample, with linear additive mode of pooled weights from both European and East-Asian schizophrenia PGC GWAS, SCZcross-ancestry-PRS_csx_ was nominally significantly associated with TRS status (OR = 1.19, 95% CI=1.02-1.38, p = 0.03), despite results from conventional PRS-CS were non-significant. In STRATA-G sample, SCZ_EUR_-PRS_cs_ and TRS-PRS_cs_ remained non-significant. The violin and quantile plots are included in **Supplementary Figures 9-12**.

In pathway-based PRS analyses, none of the TRS-pPRS calculated from TRS-PGC GWAS survived FDR correction in both the HK and STRATA-G samples. Likewise, no pathway in SCZ_EAS_-pPRS survived FDR correction in the HK sample. However, using SCZ_EUR_-pPRS, the *translation at presynapse* (OR=1.29; P_FDR_=0.05) and *presynaptic ribosome* (OR=1.30; P_FDR_ =0.05) pathways were significant in the HK sample after FDR correction. Individuals in the top pPRS quantile exhibited higher odds of TRS compared with those in the bottom quantile for the translation at presynapse pathway (OR = 1.69, p = 0.03) and the presynaptic ribosome pathway (OR = 2.27, p = 0.03). The pPRS of nominally significant pathways, associated violin and quantile plots were shown in **Supplementary Tables 6-7** and **Supplementary Figures 13-14**.

### 3.4 TWAS

After tissue-specific FDR correction, five genes across six gene-tissue pairs were significant in the HK cohort. The significant genes were ZXDC (Beta_spinal cord cervical_ = -1.5, P_tissue-FDR_ = 0.007; Beta_putamen basal ganglia_ = -1.0, P_tissue-FDR_ = 0.02), CBWD1 (Beta_nucleus accumbens basal ganglia_ = 0.6, P_tissue-FDR_ = 0.01), C9orf85 (Beta_nucleus accumbens basal ganglia_ = -2.3, P_tissue-FDR_ = 0.01), AQP7 (Beta_spinal cord cervical_ = 0.6, P_tissue-FDR_ = 0.03), and CCNDBP1 (Beta_anterior cingulate cortex_ = -1.9, P_tissue-FDR_ = 0.05). In STRATA-G, only DEK (Beta_putamen basal ganglia_ = -1.5, P_tissue-FDR_ = 0.003) showed a significant association.

Notably, all but one HK-associated gene showed evidence of cross-cohort heterogeneity (P_diff_ < 0.05), driven by reduced effect sizes in STRATA-G. Several genes also exhibited opposite effect directions between cohorts, albeit non-significant in STRATA-G. Given the sample sizes of both cohorts, these findings should be interpreted cautiously, as the data are insufficient to distinguish between true biological discordance and sampling variability. Meta-analysis of TWAS results, which tested the union of genes across cohorts (7% more than HK and 13% more than STRATA-G), did not identify any gene with tissue-FDR < 0.1. Detailed results are shown in **Table 2**.

**Table 2.**
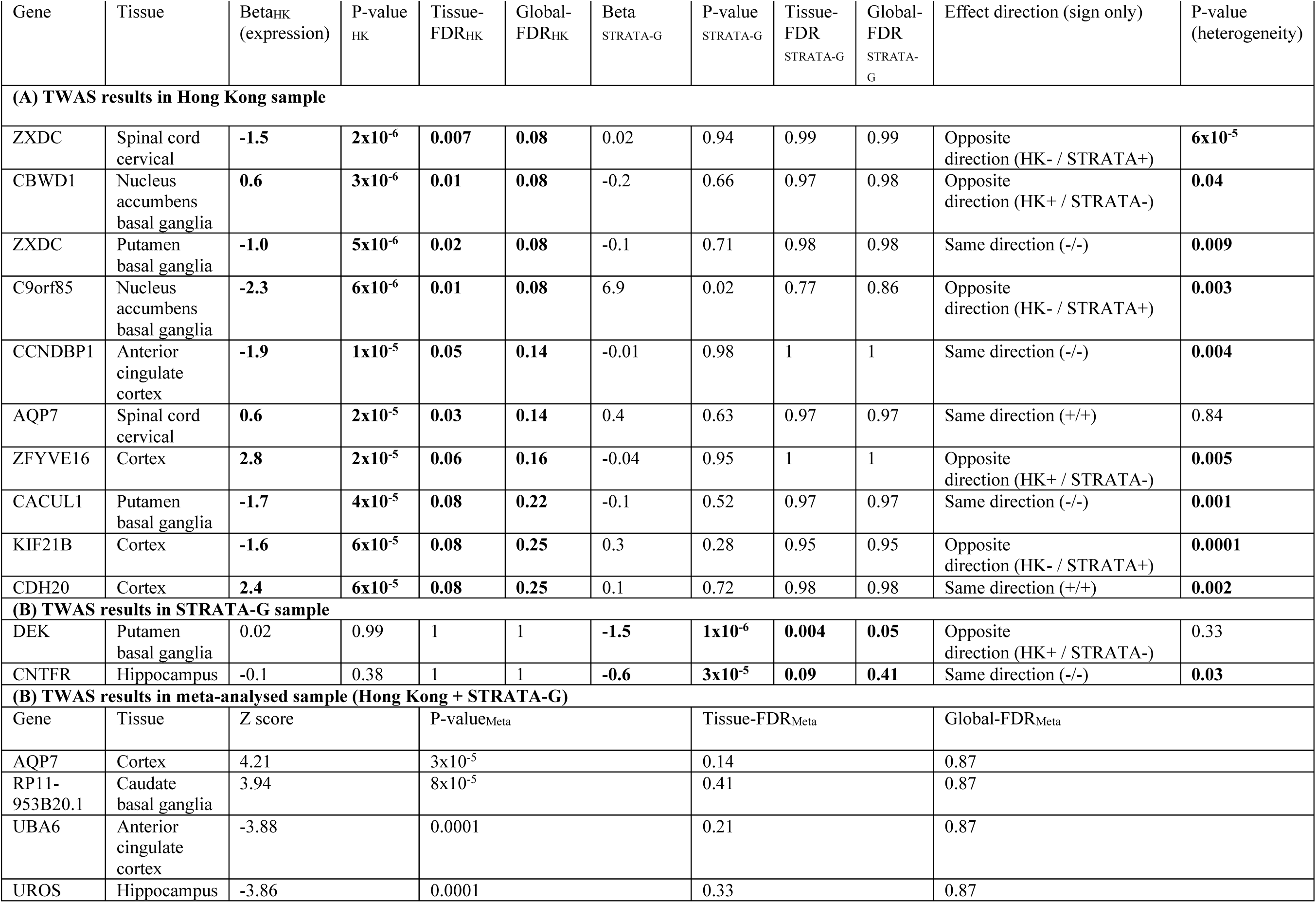
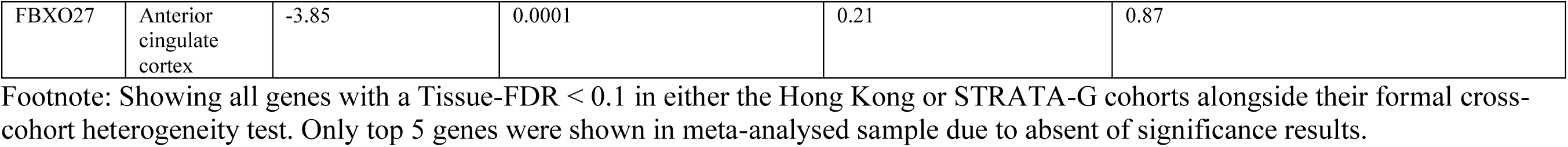
Top-ranked gene identified by TWAS in Hong Kong and STRATA-G samples.

| Gene | Tissue | Beta<sub>HK</sub><br>(expression) | P-value<sub>HK</sub> | Tissue-FDR<sub>HK</sub> | Global-FDR<sub>HK</sub> | Beta<sub>STRATA-G</sub> | P-value<sub>STRATA-G</sub> | Tissue-FDR<sub>STRATA-G</sub> | Global-FDR<sub>STRATA-G</sub> | Effect direction (sign only) | P-value (heterogeneity) |
| --- | --- | --- | --- | --- | --- | --- | --- | --- | --- | --- | --- |
| **(A) TWAS results in Hong Kong sample** |  |  |  |  |  |  |  |  |  |  |  |
| ZXDC | Spinal cord cervical | **-1.5** | **2x10<sup>-6</sup>** | **0.007** | **0.08** | 0.02 | 0.94 | 0.99 | 0.99 | Opposite direction (HK- / STRATA+) | **6x10<sup>-5</sup>** |
| CBWD1 | Nucleus accumbens basal ganglia | **0.6** | **3x10<sup>-6</sup>** | **0.01** | **0.08** | -0.2 | 0.66 | 0.97 | 0.98 | Opposite direction (HK+ / STRATA-) | **0.04** |
| ZXDC | Putamen basal ganglia | **-1.0** | **5x10<sup>-6</sup>** | **0.02** | **0.08** | -0.1 | 0.71 | 0.98 | 0.98 | Same direction (-/-) | **0.009** |
| C9orf85 | Nucleus accumbens basal ganglia | **-2.3** | **6x10<sup>-6</sup>** | **0.01** | **0.08** | 6.9 | 0.02 | 0.77 | 0.86 | Opposite direction (HK- / STRATA+) | **0.003** |
| CCNDBP1 | Anterior cingulate cortex | **-1.9** | **1x10<sup>-5</sup>** | **0.05** | **0.14** | -0.01 | 0.98 | 1 | 1 | Same direction (-/-) | **0.004** |
| AQP7 | Spinal cord cervical | **0.6** | **2x10<sup>-5</sup>** | **0.03** | **0.14** | 0.4 | 0.63 | 0.97 | 0.97 | Same direction (+/+) | 0.84 |
| ZFYVE16 | Cortex | **2.8** | **2x10<sup>-5</sup>** | **0.06** | **0.16** | -0.04 | 0.95 | 1 | 1 | Opposite direction (HK+ / STRATA-) | **0.005** |
| CACUL1 | Putamen basal ganglia | **-1.7** | **4x10<sup>-5</sup>** | **0.08** | **0.22** | -0.1 | 0.52 | 0.97 | 0.97 | Same direction (-/-) | **0.001** |
| KIF21B | Cortex | **-1.6** | **6x10<sup>-5</sup>** | **0.08** | **0.25** | 0.3 | 0.28 | 0.95 | 0.95 | Opposite direction (HK- / STRATA+) | **0.0001** |
| CDH20 | Cortex | **2.4** | **6x10<sup>-5</sup>** | **0.08** | **0.25** | 0.1 | 0.72 | 0.98 | 0.98 | Same direction (+/+) | **0.002** |
| **(B) TWAS results in STRATA-G sample** |  |  |  |  |  |  |  |  |  |  |  |
| DEK | Putamen basal ganglia | 0.02 | 0.99 | 1 | 1 | **-1.5** | **1x10<sup>-6</sup>** | **0.004** | **0.05** | Opposite direction (HK+ / STRATA-) | 0.33 |
| CNTFR | Hippocampus | -0.1 | 0.38 | 1 | 1 | **-0.6** | **3x10<sup>-5</sup>** | **0.09** | **0.41** | Same direction (-/-) | **0.03** |
| **(B) TWAS results in meta-analysed sample (Hong Kong + STRATA-G)** |  |  |  |  |  |  |  |  |  |  |  |
| Gene | Tissue | Z score |  | P-value<sub>Meta</sub> |  |  | Tissue-FDR<sub>Meta</sub> |  | Global-FDR<sub>Meta</sub> |  |  |
| AQP7 | Cortex | 4.21 |  | 3x10<sup>-5</sup> |  |  | 0.14 |  | 0.87 |  |  |
| RP11-953B20.1 | Caudate basal ganglia | 3.94 |  | 8x10<sup>-5</sup> |  |  | 0.41 |  | 0.87 |  |  |
| UBA6 | Anterior cingulate cortex | -3.88 |  | 0.0001 |  |  | 0.21 |  | 0.87 |  |  |
| UROS | Hippocampus | -3.86 |  | 0.0001 |  |  | 0.33 |  | 0.87 |  |  |

|  |  |  |  |  |  |
| --- | --- | --- | --- | --- | --- |
| FBXO27 | Anterior<br>cingulate<br>cortex | -3.85 | 0.0001 | 0.21 | 0.87 |

Footnote: Showing all genes with a Tissue-FDR < 0.1 in either the Hong Kong or STRATA-G cohorts alongside their formal cross-cohort heterogeneity test. Only top 5 genes were shown in meta-analysed sample due to absent of significance results.

### 3.5 Gene set enrichment analyses

The subsequent GSEA based on TWAS expressions identified three synaptic pathways in the Hong Kong cohort reaching tissue-level FDR < 0.1, including *postsynaptic specialization* in the hypothalamus (NES = -1.6, P_tissue-FDR_ = 0.03), *presynaptic membrane* in the putamen basal ganglia (NES = -1.8, P_tissue-FDR_ = 0.07), and *process in the presynapse* in the putamen basal ganglia (NES = -1.6, P_tissue-FDR_ = 0.09). In the STRATA-G cohort, two hippocampal pathways showed significant enrichment in the opposite direction, including *process in the postsynapse* (NES = 1.9, P_tissue-FDR_ = 0.01) and *regulation of postsynaptic membrane neurotransmitter receptor levels* (NES = 1.8, P_tissue-FDR_ = 0.05), whereas corresponding signals in the Hong Kong cohort were non-significant. These hippocampal pathways exhibited significant cross-cohort heterogeneity (P_diff_ < 0.01), driven by opposing effect directions, whereas the shared negative signs for subcortical pathways merely reflected non-significant noise in STRATA-G (P_diff_ >0.05). Meta-analysed GSEA across both cohorts did not identify any pathway reaching tissue-FDR < 0.1. Detailed results are shown in **Table 3**.

**Table 3.** Top-ranked synaptic pathways identified by GSEA from the TWAS expression signals in Hong Kong and STRATA-G samples.

| Pathway | Tissue | Number of genes (HK/STRAT A-G) | NES<sub>HK</sub> | P-value<sub>HK</sub> | Tissue-FDR<sub>HK</sub> | Global-FDR<sub>HK</sub> | NES<sub>STRATA-G</sub> | P-value<sub>STRATA-G</sub> | Tissue-FDR<sub>STRATA-G</sub> | Global-FDR<sub>STRAT A-G</sub> | Effect direction (sign only) | P-value (heterogeneity) |
| --- | --- | --- | --- | --- | --- | --- | --- | --- | --- | --- | --- | --- |
| **(A) GSEA results in Hong Kong sample** |  |  |  |  |  |  |  |  |  |  |  |  |
| Postsynaptic specialization | Hypothalamus | 17/45 | **-1.6** | **0.03** | **0.03** | **0.91** | -0.8 | 0.84 | 0.92 | 1 | Same direction (-/-) | 0.17 |
| Presynaptic membrane | Putamen basal ganglia | 30/30 | -1.8 | 0.003 | 0.07 | 0.82 | -0.8 | 0.74 | 0.98 | 0.99 | Same direction (-/-) | 0.06 |
| Process in the presynapse | Putamen basal ganglia | 60/59 | -1.6 | 0.008 | 0.09 | 0.91 | -0.8 | 0.88 | 0.98 | 1 | Same direction (-/-) | 0.08 |
| **(B) GSEA results in STRATA-G sample** |  |  |  |  |  |  |  |  |  |  |  |  |
| Process in the postsynapse | Hippocampus | 38/39 | -1.1 | 0.35 | 0.60 | 0.97 | **1.9** | **0.0006** | **0.01** | **0.15** | Opposite direction (-/+) | 0.002 |
| Regulation of postsynaptic membrane neurotransmitter receptor levels | Hippocampus | 23/25 | -1.0 | 0.39 | 0.60 | 0.97 | **1.8** | **0.006** | **0.05** | **0.70** | Opposite direction (-/+) | 0.01 |
| **(C) GSEA results in meta-analysed sample (Hong Kong + STRATA-G)** |  |  |  |  |  |  |  |  |  |  |  |  |
| Pathway | Tissue | Number of genes | NES<sub>Meta</sub> |  | P-value<sub>Meta</sub> |  | Tissue-FDR<sub>Meta</sub> |  | Global-FDR<sub>Meta</sub> |  |  |  |
| trans-synaptic signaling | Spinal cord cervical | 22 | 1.60 |  | 0.01 |  | 0.22 |  | 0.83 |  |  |  |
| Synaptic vesicle cycle | Substantia nigra | 18 | -1.60 |  | 0.02 |  | 0.14 |  | 0.83 |  |  |  |
| Process in the presynapse | Frontal cortex | 47 | 1.44 |  | 0.02 |  | 0.39 |  | 0.83 |  |  |  |
| Postsynaptic density membrane | Spinal cord cervical | 18 | 1.54 |  | 0.03 |  | 0.22 |  | 0.83 |  |  |  |
| Presynaptic membrane | Amygdala | 19 | -1.56 |  | 0.03 |  | 0.22 |  | 0.83 |  |  |  |

Footnote: Showing all genes with Tissue-FDR < 0.1 for Hong Kong and STRATA-G sample. Only top 5 genes were shown in meta-analysed sample due to absent of significance results.

### 3.6 Additional/sensitivity analyses

Additional MAGMA gene-set and GSEA analyses including broader brain-related pathways supported the predominance of synaptic signals. *Postsynaptic specialization* in the hypothalamus remained tissue-FDR significant in the HK sample (NES = -1.6, P_tissue-FDR_ = 0.05), and *processes in the postsynapse* in the hippocampus remained tissue-FDR significant in STRATA-G (NES = 1.9, P_tissue-FDR_ = 0.02) despite testing 127 additional pathways. Among nominally significant pathways, synaptic gene-sets accounted for four of six in HK and eleven of thirteen in STRATA-G, suggesting that synaptic pathways exhibited a higher concentration of enrichment signals than broader brain structural pathways, further supporting a crucial involvement of synaptic dysregulation in SSD treatment resistance. Full results are provided in **Supplementary Tables 8–9**. Meta-analysed TWAS without INT and STRATA-G analyses excluding PCA-predicted non-European individuals (n = 50) yielded largely consistent results with core analyses (**Supplementary Tables 10–13**).

## 4. Discussion

This is the first cross-ancestry study to investigate whether TRS-specific genetic signals are shared across SNP, gene, pathway, polygenic, and transcriptomic levels in East-Asian and European-ancestry cohorts. GWAS was conducted as an upstream intermediate step to generate harmonised summary statistics rather than as the primary interpretive focus, because of the limited statistical power for single-variant discovery. Nevertheless, by integrating cross-cohort GWAS with downstream aggregation approaches, we found that while single-variant and single-gene findings were largely cohort-specific, pathway-level and transcriptomic analyses revealed recurrent synaptic and neurobiological patterns. This suggests that, in clinical cohorts where single-locus replication is hindered by modest statistical power and genetic heterogeneity, pathway-level aggregation may better capture broader biological signals.

Across post-GWAS analyses, the most recurrent evidence implicated synaptic biology. MAGMA gene-set analyses highlighted enrichment of synaptic pathways, including regulation of synaptic vesicle exocytosis and neurotransmitter receptor transport, while TWAS-derived GSEA identified tissue-specific enrichment of postsynaptic pathways, including postsynaptic specialization in the hypothalamus and regulation of postsynaptic membrane neurotransmitter receptor levels in the hippocampus. The regulation of synaptic vesicle exocytosis at the presynaptic active zone is a core determinant of synaptic strength and short-term plasticity, governing the release and transmission of glutamate and GABA (Südhof, 2012). Dysregulated glutamatergic and GABAergic signalling provides a biologically plausible substrate for treatment-resistance (Murray et al., 2026). Together, these findings are consistent with involvement of both presynaptic and postsynaptic processes, including vesicle cycling, protein synthesis, and receptor regulation, which also aligns with neuroimaging evidence of limited dopaminergic abnormalities in TRS compared to treatment-responsive SSD (Demjaha et al., 2012). However, because individual genes and loci did not replicate across cohorts, these cohort-specific findings should be interpreted as broad biological evidence prioritizing candidate pathways rather than definitive TRS risk loci. It remains challenging to distinguish population-specific biology from sampling variability at the single-variant level.

Furthermore, while many implicated genes map to loci previously associated with schizophrenia susceptibility, our results offer a novel contribution by extending prior knowledge to suggest specific associations with TRS liability in SSD. The tissue-specific TWAS signals mapped to key structures within the medial frontal region (anterior cingulate cortex) alongside the striatal region (putamen and nucleus accumbens basal ganglia) and limbic structure (hippocampus, hypothalamus) (Ernst & Fudge, 2009; Torrico & Abdijadid, 2026), consistent with the current imaging evidence that TRS has abnormal frontal-striatal and limbic circuitry relative to non-TRS patients (Ochi et al., 2020; Zhang et al., 2025). At the gene level, several identified candidates align with established neurodevelopmental and psychiatric pathways. The *DEK* gene is associated with the brain pathways of DNA repair, cell proliferation, apoptosis, and inflammatory responses in human brain, as well as the neural development-relevant pathways (e.g., synaptic transmission, neurite outgrowth and myelination) (Greene et al., 2022). A postmortem brain tissue study linked *DEK* expression in the anterior cingulate cortex to cognitive decline in elderly patients with schizophrenia (O’Donovan et al., 2018). The *OSBPL9* gene, found significant in MAGMA in our HK sample, was previously reported as upregulated in schizophrenia organoids (Notaras et al., 2021). *ZXDC* is a co-activator of Major Histocompatibility Complex (MHC) class II gene transcription (Ma’ayan Laboratory of Computational Systems Biology, 2026), and MHC is one of the strongest population-level genetic risk loci in schizophrenia architecture (Sekar et al., 2016).

Our study has several notable strengths. Previous genetic studies in Taiwanese Han Chinese populations examined TRS by contrasting cases with non-patient controls (Liou et al., 2012), while a more recent Singaporean Chinese study compared TRS with treatment-responsive schizophrenia (Lim et al., 2023). Building on these works, the present study represents the largest GWAS to-date, by number of TRS cases, to examine treatment-resistance through direct comparison between TRS and treatment-responsive schizophrenia within a Chinese population. Although some individual STRATA-G cohorts have previously been used in pathway-specific PRS analyses of psychosis patients and healthy controls (Pistis et al., 2022), our study extends to treatment resistance among patients with TRS. The inclusion of both Chinese and European-ancestry samples enabled cross-ancestry comparison of population-specific heterogeneity and shared pathway-level patterns, supporting prior evidence that multi-ancestry studies are essential for characterising complex psychiatric phenotypes (Zhang et al., 2023). For example, the top SNP in the STRATA-G sample, rs79478186, has a MAF of 0.0008 in East Asian populations, whereas rs138175807, the top hit in the HK sample, has a MAF of 0.003 in European populations in gnomAD v4 (National Center for Biotechnology Information, 2026a, 2026b). Such large disparities in MAF highlight how population-specific genetic architectures can drive distinct downstream signals in post-GWAS analyses. Moreover, although PRS derived from large European GWAS showed significant associations in the HK sample, reflecting shared common-variant risk, PRS alone provided limited mechanistic insight. We therefore complemented PRS with TWAS to test associations with predicted gene expression and obtain directional insights into putative regulatory mechanisms across brain tissues, allowing us to explore potential convergence and divergence of biological pathways across ancestries. The use of individual-level genotype data for TWAS preserved allelic and LD structure, and enabled cohort-specific covariate adjustment, strengthening the interpretability of predicted gene-expression effects. Stringent tissue-specific FDR correction was prioritised to reflect the tissue-dependent nature of gene expression and avoid over-penalisation of biologically meaningful, tissue-restricted signals. Meta- and sensitivity analyses further increased the multiple-testing burden by evaluating the union of genes and gene-sets across cohorts, representing a conservative analytic strategy that prioritised robustness.

Nevertheless, this study has limitations. First, both the HK and STRATA-G samples were underpowered due to modest sample sizes and substantial clinical heterogeneity among patients with TRS (Demjaha et al., 2017; Lally et al., 2016). Limited power constrained replication at the SNP and gene levels and contributed to imprecision in effect estimates, plausibly contributing to the inconsistent effect directions across cohorts. Moreover, the HK sample was restricted to SSD, whereas STRATA-G included non-SSD psychoses. Although TRS is defined by treatment-response rather than baseline diagnosis, this mismatch in the two cohorts may have introduced additional variance, affecting GWAS discovery and downstream comparability. While the broader STRATA-G inclusion reflects real-world clinical heterogeneity, it may also capture variability related to illness trajectory, treatment adherence, and social vulnerability, further complicating interpretation. Secondly, TRS ascertainment relied mainly on clozapine prescription, which may introduce misclassification because clozapine may infrequently be prescribed for severe tardive dyskinesia. This proxy definition may result in the signals observed in motor-related cervical spinal cord tissue. Different TRS definitions across cohorts also reflect real-world clinical practice. The retrospectively collected HK sample relied on clozapine proxy after at least two other antipsychotic trials, whereas the prospective STRATA-G cohort applied a broader definition permitting either clozapine initiation or persistent symptoms despite adequate treatments, its data were collected before the TRS consensus definition was established. Third, although multiple strategies were used to account for population stratification across ancestries, including PRS-CSx and combined-reference clumping in GWAS meta-analysis, differences in allele frequencies and LD structure across populations cannot be fully eliminated. Incomplete phenotypic data also limited covariate adjustment and may leave residual confounding. For example, ancestry information was not documented in STRATA-G, although sensitivity analyses excluding PCA-defined non-European individuals did not alter the results. Fourth, TWAS findings should be interpreted cautiously. Because TWAS relies on eQTL-predicted gene expression, LD hitchhiking can reduce its specificity, as LD between predictive eQTLs and causal GWAS variants can yield spurious associations that tag the correct locus but implicate the wrong gene (Okamoto et al., 2023). While TWAS alongside locus-level colocalization can resolve these LD-driven signals with high specificity with reduced sensitivity (Hukku et al., 2022), these data are not currently available in our cohorts. Moreover, predicted gene-expression models may have limited generalisability across ancestries, cell types, and environmental contexts, as GTEx v8 is predominantly based on European-ancestry samples. Finally, gene-set analyses can be biologically confounded by overlapping genes, whereby pathways may be spuriously correlated with TRS due to shared genes with other causal gene-sets rather than independent biological contributions (de Leeuw et al., 2018). Our Jaccard similarity network partly addressed this issue and indicated that most synaptic pathway signals were not driven by excessive gene overlap.

In conclusion, this study provides triangulating evidence that aberrant synaptic regulatory pathways contribute to the neurobiology of TRS liability in SSD. These synaptic processes may represent a biologically coherent component of TRS liability beyond genome-wide schizophrenia risk, possibly explaining the limited treatment-response to D2-blocking agents in TRS patients. Furthermore, the enrichment of synaptic regulatory pathways within frontostriatal and limbic regions directly links TRS to localised neural circuit dysfunction. By integrating multi-level analyses, we demonstrate how pathway-level approaches may resolve convergent mechanisms even when single-variant discovery is underpowered. These findings together highlight synaptic processes within key neural circuits as a core biological substrate underlying treatment-resistance, pointing toward therapeutic targets beyond traditional dopaminergic blockade. Future research integrating fine-mapping, colocalised TWAS signals, and functional validation will be critical to translate these synaptic signals into actionable therapeutic targets.

## Data Availability

The data that support the findings of this study are available from the corresponding author upon reasonable request.

## Declaration of Interests

All authors declare no competing interests.

## Funding Disclosures

This study was funded by the HKU Seed Fund for Basic Research for New Staff (no. 202009185071) and the HKU Enhanced Start-up Fund for New Staff, granted to S.S.Y.L. This study was also supported partially by a Health and Medical Research Fund (no. 07180376) and the Lo Kwee Seong Biomedical Research Fund from The Chinese University of Hong Kong, granted to H.C.S.

## Acknowledgments

The authors thank the Information Technology Services at the University of Hong Kong for providing access to HKU research computing facilities (HPC) and the King’s College London e-Research for providing access to King’s Computational Research, Engineering and Technology Environment (CREATE).

We acknowledge other members who were involved in and contributed to the STRATA consortium, including [AESOP] Arsime Demjaha, [Belfast] Lina Homman, Domenico Berardi, [Bologna] Giuseppe D’Andrea, Elena Bonora, Lorenzo Guidi, Ornella Lastrina, Roberto Muratori [GAP] Olesya Ajnakina, [Istanbul] Handan Noyan, [Lausanne] Sara Camporesi, Martine Cleusix, Raoul Jenni, Ines Khadimallah, Romeo Restellini, [Oslo] Carmen Simonsen, [Paris] Aziz Ferchiou, Jean-Romain Richard, Andrei Szöke, [STRATA-G] Deborah Agbedjro, Laura Kassoumeri, Daniel Stahl, Michael O’Donovan.

## Declaration of Generative AI and AI-assisted technologies in the writing process

During the preparation of this work, Perry BM Leung used ChatGPT to assist with validation of statistical code written in R and with language editing, including spell checks and improving the clarity of explanations of statistical concepts. After using this tool, the authors reviewed and edited the content as needed and take full responsibility for the accuracy and integrity of the work.

## Notes

### Competing Interest Statement

The authors have declared no competing interest.

### Author Declarations

This study was approved by the institutional review boards of the Hong Kong Hospital Authority and participating hospitals (CPH: NTWC/CREC/823/10, NTWC/CREC/1293/14, CIRB-2023-025-4; QMH: UW-22-724), and written informed consent was obtained from all participants.

### Summary of Updates

The manuscript has been revised to update author affiliations and correct several grammatical/typographical errors throughout the text.

