## Supplementary Materials for "Genetic architecture of treatment-resistant schizophrenia across East Asian and European cohorts: insights from GWAS, TWAS, and synaptic pathway analyses"

### Supplementary Table 1. Clinical characteristics of Hong Kong sample included in the analysis

|  | **TRS (n=290)** | **Non-TRS (n=508)** | **P-value** |
| --- | --- | --- | --- |
| Male (%) | 156 (53.8%) | 222 (43.7%) | 0.008 |
| Onset age (median) | 20.6 (19) | 27.4 (25) | <2x10^-16^ |
| Current age (31/08/2025-DOB) (median) | 39.0 (37.4) | 41.4 (39.7) | 0.0002 |
| Self-reported psychiatric family history (%) | 118 (40.7%) | 126 (24.8%) | 8x10^-14^ |
| History of self-harm (%) | 99 (34.1%) | 86 (16.9%) | 5x10^-8^ |
| History of aggressive behaviour (%) | 97 (33.4%) | 89 (17.5%) | 1x10^-6^ |
| Years of education (median) | 11.42 (11.0) | 11.75 (11.0) | 0.10 |
| Immigrantion status (born in Mainland China or overseas) (%) | 86 (29.7%) | 167 (32.9%) | 0.66 |
|  | **Retrospective clozapine cohort (n=175)** | **FEP cohort (n=623)** | **P-value** |
| Male (%) | 84 (48%) | 294 (47%) | 0.92 |
| Onset age (median) | 20.4 (19) | 26.1 (24) | <2x10^-16^ |
| Current age (31/08/2025-DOB) (median) | 40.6 (41.0) | 40.5(38.5) | 0.86 |
| Self-reported psychiatric family history (%) | 76 (43.4%) | 168 (27.0%) | 5x10^-8^ |
| History of self-harm (%) | 64 (37%) | 121 (19.4%) | 2x10^-5^ |
| History of aggressive behaviour (%) | 56 (32%) | 130 (20.9%) | 0.005 |
| Years of education (median) | 11.4 (11.0) | 11.7 (11.0) | 0.24 |
| Immigrantion status (born in Mainland China or overseas) (%) | 49 (28%) | 204 (33%) | 0.27 |

### Supplementary Table 2. Clinical characteristics of STRATA-G sample included in the analysis (with missingness shown)

|  | Missingness | **TRS (n=168)** | **Non-TRS (n=1075)** | **P-value** |
| --- | --- | --- | --- | --- |
| Male (%) | 0% | 118 (70.2%) | 608 (56.6%) | 0.001 |
| Onset age (median) | 15.2% | 23.9 (21.5) | 27.3 (25.1) | 2x10^-6^ |
| Current age (31/08/2025-DOB) (median) | 36.9% | 45.5 (43.3) | 48.8 (46.9) | 0.001 |
| Duration of untreated psychosis (days) | 29.0% | 186.6 (30.4) | 318.7 (24.0) | 0.03 |

### Supplementary Table 3. Cohorts included in STRATA-G

| Cohort (Country) | Number of subjects with TRS | Number of subjects with non-TRS |
| --- | --- | --- |
| AESOP (UK) | 17 | 37 |
| Belfast (UK) | 13 | 83 |
| Bologna (Italy) | 2 | 34 |
| GAP (UK) | 30 | 125 |
| Istanbul (Turkey) | 17 | 26 |
| Lausanne (Switzerland) | 18 | 180 |
| Oslo (Norway) | 3 | 117 |
| Paris (France) | 6 | 25 |
| Prague (Czech) | 9 | 34 |
| Santander (Spain) | 47 | 308 |
| Sao Paulo (Brazil) | 2 | 41 |
| UCL (UK) | 4 | 65 |

### Supplementary Table 4. Lead SNP identified by GWAS in Hong Kong and STRATA-G samples

| SNP | CHR | BP | A1/A2 | MAF | OR | SE | P |
| --- | --- | --- | --- | --- | --- | --- | --- |
| **(A) GWAS results in Hong Kong sample** | | | | | | | |
| **rs138175807** | 15 | 22746909 | A/G | 0.48 | 1.69 | 0.11 | 1x10^-6^ |
| **rs75647477** | 8 | 93352764 | A/G | 0.07 | 0.27 | 0.28 | 3x10^-6^ |
| **rs3860448** | 2 | 105989649 | A/G | 0.24 | 0.54 | 0.13 | 4x10^-6^ |
| **rs55772829** | 5 | 23727984 | T/C | 0.37 | 1.64 | 0.11 | 6x10^-6^ |
| rs6543355 | 2 | 105976195 | G/T | 0.76 | 0.55 | 0.13 | 7x10^-6^ |
| rs1669510 | 2 | 106005140 | A/G | 0.73 | 0.57 | 0.12 | 8x10^-6^ |
| **rs10570030** | 10 | 65258703 | A/AACAC | 0.13 | 2.00 | 0.16 | 9x10^-6^ |
| **rs2244476** | 8 | 14742881 | A/G | 0.30 | 0.58 | 0.12 | 1x10^-5^ |
| **(B) GWAS results in Strata-G sample** | | | | | | | |
| **rs79478186** | 5 | 113189975 | C/G | 0.04 | 3.61 | 0.26 | 6x10^-7^ |
| **rs11207645** | 1 | 60669675 | A/G | 0.02 | 5.80 | 0.36 | 1x10^-6^ |
| **rs35258147** | 3 | 117010426 | T/C | 0.22 | 1.96 | 0.14 | 2x10^-6^ |
| rs10460824 | 3 | 116994873 | A/G | 0.22 | 1.91 | 0.14 | 5x10^-6^ |
| **rs62559696** | 9 | 28574932 | C/T | 0.02 | 4.47 | 0.33 | 5x10^-6^ |
| **rs78532950** | 1 | 21592253 | G/A | 0.03 | 3.62 | 0.29 | 9x10^-6^ |
| **rs11794329** | 9 | 13345643 | A/C | 0.24 | 0.47 | 0.17 | 9x10^-6^ |
| rs76782877 | 1 | 21594400 | T/C | 0.03 | 3.61 | 0.29 | 1x10^-5^ |
| **(C) GWAS results in meta-analysed sample (Hong Kong + STRATA-G)** | | | | | | | |
| **rs138175807** | 15 | 22746909 | A/G | NA | 1.69 | 0.18 | 1x10^-6^ |
| **rs76592182** | 9 | 115725251 | T/C | NA | 0.41 | 0.08 | 1x10^-6^ |
| **rs78947414** | 11 | 69146940 | T/C | NA | 2.04 | 0.31 | 2x10^-6^ |
| [rs76146833](https://www.ncbi.nlm.nih.gov/snp/rs76146833) | 11 | 69148569 | T/C | NA | 1.98 | 0.29 | 2x10^-6^ |
| [rs79580557](https://www.ncbi.nlm.nih.gov/snp/rs79580557) | 9 | 115719377 | A/G | NA | 2.45 | 0.47 | 3x10^-6^ |
| [rs76196028](https://www.ncbi.nlm.nih.gov/snp/rs76196028) | 11 | 69141754 | A/G | NA | 0.51 | 0.07 | 3x10^-6^ |
| **rs75647477** | 8 | 93352764 | A/G | NA | 0.27 | 0.08 | 3x10^-6^ |
| [rs60659361](https://www.ncbi.nlm.nih.gov/snp/rs60659361) | 11 | 69140554 | T/C | NA | 1.96 | 0.29 | 3x10^-6^ |
| [rs7045079](https://www.ncbi.nlm.nih.gov/snp/rs7045079) | 9 | 115723485 | T/C | NA | 2.39 | 0.45 | 4x10^-6^ |
| [rs11228525](https://www.ncbi.nlm.nih.gov/snp/rs11228525) | 11 | 69148203 | T/C | NA | 1.95 | 0.28 | 4x10^-6^ |
| [rs11228524](https://www.ncbi.nlm.nih.gov/snp/rs11228524) | 11 | 69148200 | A/G | NA | 0.51 | 0.07 | 4x10^-6^ |
| [rs12222313](https://www.ncbi.nlm.nih.gov/snp/rs12222313) | 11 | 69151839 | T/C | NA | 0.49 | 0.08 | 4x10^-6^ |
| [rs11228522](https://www.ncbi.nlm.nih.gov/snp/rs11228522) | 11 | 69148063 | T/C | NA | 1.92 | 0.27 | 4x10^-6^ |
| r[s74895111](https://www.ncbi.nlm.nih.gov/snp/rs74895111) | 11 | 69141980 | A/C | NA | 0.52 | 0.07 | 5x10^-6^ |
| [rs79196642](https://www.ncbi.nlm.nih.gov/snp/rs79196642) | 11 | 69147244 | T/C | NA | 1.94 | 0.28 | 5x10^-6^ |
| [rs11228530](https://www.ncbi.nlm.nih.gov/snp/rs11228530) | 11 | 69151517 | A/C | NA | 0.52 | 0.07 | 5x10^-6^ |
| [rs11228528](https://www.ncbi.nlm.nih.gov/snp/rs11228528) | 11 | 69150704 | T/C | NA | 0.51 | 0.08 | 6x10^-6^ |
| [rs74308250](https://www.ncbi.nlm.nih.gov/snp/rs74308250) | 9 | 115737123 | T/C | NA | 0.42 | 0.08 | 6x10^-6^ |
| **rs55772829** | 5 | 23727984 | T/C | NA | 1.64 | 0.18 | 6x10^-6^ |
| **rs7906005** | 10 | 120057378 | T/C | NA | 1.79 | 0.23 | 7x10^-6^ |
| **rs6543355** | 2 | 105976195 | T/G | NA | 0.55 | 0.07 | 7x10^-6^ |
| [rs117811876](https://www.ncbi.nlm.nih.gov/snp/rs117811876) | 11 | 69149883 | T/C | NA | 0.52 | 0.08 | 8x10^-6^ |
| [rs75537789](https://www.ncbi.nlm.nih.gov/snp/rs75537789) | 9 | 115711813 | A/G | NA | 2.28 | 0.42 | 8x10^-6^ |
| [rs1669510](https://www.ncbi.nlm.nih.gov/snp/rs1669510) | 2 | 106005140 | A/G | NA | 1.76 | 0.22 | 8x10^-6^ |
| [rs11228527](https://www.ncbi.nlm.nih.gov/snp/rs11228527) | 11 | 69149614 | A/G | NA | 0.53 | 0.08 | 8x10^-6^ |
| **rs76740325** | 5 | 158627795 | T/G | NA | 0.55 | 0.07 | 9x10^-6^ |
| [rs74674916](https://www.ncbi.nlm.nih.gov/snp/rs74674916) | 5 | 158627931 | A/C | NA | 1.81 | 0.24 | 1x10^-5^ |
| [rs12773384](https://www.ncbi.nlm.nih.gov/snp/rs12773384) | 10 | 120051901 | A/G | NA | 0.56 | 0.07 | 1x10^-5^ |

Footnote: Bolded for index SNPs after clumping, showing all variants with p<1e-5; Standard errors in meta-analysis were estimated via the delta method as exp(β) × SE(β)

### Supplementary Table 5. Top-ranked gene identified by MAGMA gene-based analysis in Hong Kong and STRATA-G samples

| Gene | CHR | START | STOP | NSNPS | P-value (MAGMA P_multi) | FDR-P |
| --- | --- | --- | --- | --- | --- | --- |
| **(A) MAGMA gene-based analysis results in Hong Kong sample** | | | | | | |
| OSBPL9 | 1 | 51581874 | 51799219 | 84 | 3x10^-6^ | 0.05 |
| KITLG | 12 | 88482793 | 88615473 | 112 | 9x10^-5^ | 0.66 |
| LOC162038 | 16 | 27031898 | 27079166 | 105 | 0.0001 | 0.66 |
| GARS1 | 7 | 30559565 | 30644033 | 88 | 0.0002 | 0.66 |
| CNMD | 13 | 52693264 | 52774812 | 95 | 0.0004 | 0.66 |
| **(B) MAGMA gene-based analysis results in STRATA-G sample** | | | | | | |
| MAMDC2 | 9 | 70008519 | 70236972 | 324 | 0.0001 | 0.56 |
| UBTD2 | 5 | 172199645 | 172318791 | 113 | 0.0001 | 0.56 |
| KRTAP5-11 | 11 | 71571855 | 71617875 | 9 | 0.0002 | 0.56 |
| TBCEL | 11 | 120989094 | 121099647 | 72 | 0.0002 | 0.56 |
| HK1 | 10 | 69234984 | 69411882 | 115 | 0.0002 | 0.56 |
| **(C) MAGMA gene-based analysis results in meta-analysed sample (Hong Kong + STRATA-G)** | | | | | | |
| TBCEL | 11 | 120989094 | 121099647 | 90 | 5x10^-6^ | 0.09 |
| AVL9 | 7 | 32460426 | 32598741 | 344 | 4x10^-5^ | 0.25 |
| IFITM10 | 11 | 1722410 | 1785594 | 84 | 4x10^-5^ | 0.25 |
| CTSD | 11 | 1742752 | 1798992 | 70 | 6x10^-5^ | 0.25 |
| LSM5 | 7 | 32475333 | 32530258 | 140 | 7x10^-5^ | 0.25 |

Footnote: only top 5 genes were shown due to absent of significance results (NSNPS<5 were excluded)

### Supplementary Table 6. Top-ranked synaptic pathways identified by pPRS in Hong Kong sample

| Pathway | Number of SNP | PRS.R2 | OR | P | FDR-P |
| --- | --- | --- | --- | --- | --- |
| **(A) SCZ_EUR_-pPRS** | | | | | |
| Translation at presynapse | 115 | 0.02 | 1.30 | 0.0009 | 0.05 |
| Presynaptic ribosome | 117 | 0.02 | 1.29 | 0.0009 | 0.05 |
| Synapse organization | 6045 | 0.01 | 1.24 | 0.006 | 0.17 |
| Translation at synapse | 143 | 0.01 | 1.23 | 0.008 | 0.17 |
| Translation at postsynapse | 143 | 0.01 | 1.23 | 0.008 | 0.17 |
| Regulation of synapse assembly | 1413 | 0.01 | 1.21 | 0.01 | 0.20 |
| Regulation of synaptic vesicle cycle | 166 | 0.009 | 1.19 | 0.02 | 0.31 |
| Regulation of presynapse assembly | 386 | 0.008 | 1.18 | 0.03 | 0.40 |
| Postsynaptic ribosome | 163 | 0.007 | 1.18 | 0.03 | 0.40 |
| Regulation protein catabolic process at postsynapse | 94 | 0.007 | 0.86 | 0.04 | 0.44 |
| Synapse assembly | 2402 | 0.006 | 1.16 | 0.05 | 0.45 |
| **(B) SCZ_EAS_-pPRS** | | | | | |
| Process in the postsynapse | 2947 | 0.009 | 1.20 | 0.02 | 0.62 |
| Structural constituent of synapse | 670 | 0.008 | 1.19 | 0.02 | 0.62 |
| Translation at presynapse | 124 | 0.007 | 1.17 | 0.04 | 0.62 |
| Presynaptic ribosome | 126 | 0.007 | 1.17 | 0.04 | 0.62 |
| Regulation of synapse assembly | 1182 | 0.006 | 1.16 | 0.05 | 0.62 |
| Regulation of postsynapse assembly | 596 | 0.006 | 1.17 | 0.05 | 0.62 |
| Postsynaptic cytoskeleton | 206 | 0.006 | 1.17 | 0.05 | 0.62 |
| Presynaptic active zone cytoplasmic component | 342 | 0.006 | 1.16 | 0.05 | 0.62 |
| **(C) TRS-pPRS** |  |  |  |  |  |
| Modulation of chemical synaptic transmission | 959 | 0.008 | 0.84 | 0.02 | 0.98 |
| Structural constituent of postsynapse | 419 | 0.008 | 0.85 | 0.03 | 0.98 |

Footnote: Showing all pathways with nominal p-value<0.05

### Supplementary Table 7. Top-ranked synaptic pathways identified by pPRS in STRATA-G sample

| Pathway | Number of SNP | PRS.R2 | OR | P | FDR-P |
| --- | --- | --- | --- | --- | --- |
| **(A) SCZ_EUR_-pPRS** | | | | | |
| Regulation of presynapse assembly | 293 | 0.01 | 0.77 | 0.004 | 0.42 |
| Synaptic vesicle exocytosis | 804 | 0.009 | 0.80 | 0.01 | 0.52 |
| Process in the postsynapse | 2983 | 0.008 | 1.24 | 0.01 | 0.52 |
| Postsynaptic neurotransmitter receptor endocytosis | 405 | 0.006 | 1.22 | 0.03 | 0.61 |
| Postsynaptic specialization assembly | 637 | 0.006 | 1.21 | 0.04 | 0.61 |
| Regulation of postsynaptic membrane neurotransmitter receptor levels | 1870 | 0.006 | 1.20 | 0.05 | 0.61 |
| **(B) TRS-pPRS** | | | | | |
| postsynaptic_recycling_endosome | 33 | 0.01 | 1.29 | 0.007 | 0.62 |
| presynaptic_active_zone | 825 | 0.006 | 1.21 | 0.04 | 0.62 |
| modification_of_synaptic_structure | 315 | 0.006 | 1.20 | 0.04 | 0.62 |
| Presynaptic active zone membrane | 588 | 0.006 | 1.20 | 0.04 | 0.62 |

Footnote: Showing all pathways with nominal p-value<0.05

### Supplementary Table 8. Top-ranked broader brain pathways identified GSEA sensitivity analysis in Hong Kong sample

| Pathway | Tissue | Number of genes | NES | P-value | Tissue-FDR | Global-FDR |
| --- | --- | --- | --- | --- | --- | --- |
| **GOBP neuron projection extension** | Putamen basal ganglia | 20 | 2.0 | 0.0006 | 0.02 | 0.24 |
| Presynaptic membrane | Putamen basal ganglia | 30 | -1.8 | 0.004 | 0.13 | 0.99 |
| Process in the presynapse | Putamen basal ganglia | 60 | -1.6 | 0.02 | 0.23 | 0.99 |
| GOBP neuron death | Cerebellar Hemisphere | 64 | -1.6 | 0.01 | 0.65 | 0.99 |
| Integral component of presynaptic membrane | Putamen basal ganglia | 24 | -1.7 | 0.01 | 0.14 | 0.99 |
| GOBP ensheathment of neurons | Frontal Cortex | 16 | 1.7 | 0.01 | 0.53 | 0.99 |
| GOBP neurotransmitter transport | Putamen basal ganglia | 62 | -1.5 | 0.03 | 0.65 | 0.99 |
| Synapse organization | Anterior cingulate cortex | 45 | -1.5 | 0.04 | 0.79 | 0.99 |
| GOBP regulation of axonogenesis | Putamen basal ganglia | 17 | 1.6 | 0.03 | 0.28 | 0.99 |
| GOBP neuron death | Spinal cord cervical | 35 | 1.6 | 0.03 | 0.36 | 0.99 |
| GOBP positive regulation of neuron death | Cerebellar Hemisphere | 18 | -1.6 | 0.03 | 0.71 | 0.99 |
| GOBP neuron projection extension | Caudate basal ganglia | 26 | 1.5 | 0.03 | 0.97 | 0.99 |
| GOBP neuron projection guidance | Frontal Cortex | 29 | 1.5 | 0.03 | 0.58 | 0.99 |
| GOBP neurotransmitter transport | Putamen basal ganglia | 31 | -1.5 | 0.04 | 0.33 | 0.99 |
| Synapse assembly | Cerebellum | 41 | 1.5 | 0.04 | 0.98 | 0.99 |
| GBOP neuron apoptotic process | Spinal cord cervical | 21 | 1.6 | 0.05 | 0.37 | 0.99 |
| Postsynaptic specialization | Hypothalamus | 17 | -1.6 | 0.05 | 0.05 | 0.99 |
| Synaptic vesicle membrane | Nucleus accumbens basal ganglia | 18 | -1.5 | 0.05 | 0.78 | 0.99 |

Footnote: Showing all pathways with nominal p-value<0.05; bolded for Tissue-FDR<0.05

### Supplementary Table 9. Top-ranked broader brain pathways identified GSEA sensitivity analysis in STRATA-G sample

| Pathway | Tissue | Number of genes | NES | P-value | Tissue-FDR | Global-FDR |
| --- | --- | --- | --- | --- | --- | --- |
| **Process in the postsynapse** | Hippocampus | 39 | 1.9 | 0.0005 | 0.02 | 0.33 |
| GOBP neuron projection guidance | Amygdala | 19 | -1.8 | 0.004 | 0.07 | 1.00 |
| Regulation of postsynaptic membrane neurotransmitter receptor levels | Hippocampus | 25 | 1.7 | 0.01 | 0.18 | 1.00 |
| GOBP positive regulation of neuron projection | Putamen basal ganglia | 28 | -1.6 | 0.02 | 0.89 | 1.00 |
| Synaptic vesicle cycle | Hippocampus | 25 | -1.6 | 0.02 | 0.29 | 1.00 |
| Synaptic vesicle | Caudate basal ganglia | 30 | -1.5 | 0.03 | 0.75 | 1.00 |
| Synaptic vesicle cycle | Cerebellar hemisphere | 51 | 1.5 | 0.04 | 0.64 | 1.00 |
| Regulation of postsynaptic membrane neurotransmitter levels | Anterior cingulate cortex | 23 | 1.5 | 0.04 | 0.57 | 1.00 |
| Synaptic vesicle exocytosis | Cerebellar hemisphere | 17 | 1.6 | 0.04 | 0.64 | 1.00 |
| GOBP dendrite development | Caudate basal ganglia | 45 | -1.4 | 0.04 | 0.75 | 1.00 |
| GOBP positive regulation of neurogenesis | Amygdala | 19 | -1.5 | 0.04 | 0.27 | 1.00 |
| GOBP regulation of neurogenesis | Amygdala | 28 | -1.5 | 0.04 | 0.27 | 1.00 |
| GOBP neurotransmitter transport | Cortex | 39 | -1.5 | 0.04 | 0.97 | 1.00 |
| Postsynaptic specialization | Caudate basal ganglia | 75 | -1.4 | 0.04 | 0.75 | 1.00 |
| Integral component of presynaptic membrane | Cerebellar hemisphere | 36 | 1.5 | 0.05 | 0.64 | 1.00 |
| Synaptic vesicle membrane | Cerebellar hemisphere | 30 | 1.5 | 0.05 | 0.64 | 1.00 |
| Chemical synaptic transmission | Cortex | 27 | -1.5 | 0.05 | 0.98 | 1.00 |

Footnote: Showing all pathways with nominal p-value<0.05; bolded for Tissue-FDR<0.05

### Supplementary Table 10. Top-ranked synaptic pathways identified by GSEA from the TWAS expression signals in sensitivity analysis of meta-analysed sample (Hong Kong and STRATA-G) (without inverse normal transformation)

| Pathway | Tissue | Number of genes | NES | P-value | Tissue-FDR | Global-FDR |
| --- | --- | --- | --- | --- | --- | --- |
| Synapse assembly | Cerebellum | 41 | 1.5 | 0.02 | 0.97 | 0.85 |
| Synaptic vesicle cycle | Substantia nigra | 18 | -1.6 | 0.03 | 0.23 | 0.85 |
| Modulation of chemical synaptic transmission | Nucleus accumbent basal ganglia | 15 | -1.6 | 0.03 | 0.34 | 0.85 |
| Synapse assembly | Cerebellar Hemisphere | 41 | 1.4 | 0.03 | 0.98 | 0.85 |
| Chemical synaptic transmission | Cortex | 31 | -1.6 | 0.04 | 0.50 | 0.85 |

Footnote: only top 5 pathways were shown due to no significant

### Supplementary Table 11. Lead SNP identified by GWAS in sensitivity analysis of STRATA-G cohort excluding PCA predicted non-European subjects

| SNP | CHR | BP | A1/A2 | MAF | OR | SE | P |
| --- | --- | --- | --- | --- | --- | --- | --- |
| rs79478186 | 5 | 113189975 | C/G | 0.04 | 3.63 | 0.26 | 5x10^-7^ |
| rs11207645 | 1 | 60669675 | A/G | 0.02 | 5.80 | 0.36 | 1x10^-6^ |
| rs35258147 | 3 | 117010426 | T/C | 0.22 | 1.96 | 0.14 | 2x10^-6^ |
| rs10460824 | 3 | 116994873 | A/G | 0.22 | 1.92 | 0.14 | 5x10^-6^ |
| rs62559696 | 9 | 28574932 | C/T | 0.02 | 4.35 | 0.33 | 7x10^-6^ |
| rs78532950 | 1 | 21592253 | G/A | 0.03 | 3.68 | 0.29 | 8x10^-6^ |
| rs76782877 | 1 | 21594400 | T/C | 0.03 | 3.67 | 0.29 | 8x10^-6^ |

Footnote: Bolded for index SNPs after clumping, showing all variants with p<1e-5

### Supplementary Table 12. Top-ranked gene identified by MAGMA gene-based analysis in sensitivity analysis of STRATA-G sample excluding PCA predicted non-European subjects

| Geneset | Number of genes | Beta | P | FDR-P |
| --- | --- | --- | --- | --- |
| Regulation of synaptic vesicle exocytosis | 33 | 0.56 | 9x10^-5^ | 0.008 |
| Synaptic vesicle cycle | 202 | 0.11 | 0.02 | 0.62 |
| Synaptic vesicle exocytosis | 75 | 0.19 | 0.02 | 0.62 |
| Process in the presynapse | 288 | 0.09 | 0.03 | 0.62 |
| Neuronal dense core vesicle | 39 | 0.22 | 0.04 | 0.66 |
| Regulation of postsynapse assembly | 56 | 0.19 | 0.04 | 0.66 |

Footnote: Showing all pathways with nominal p-value<0.05

### Supplementary Table 13. Top-ranked synaptic pathways identified by GSEA from the TWAS expression signals in sensitivity analysis of the STRATA-G sample excluding PCA predicted non-European subjects

| Pathway | Tissue | Number of genes | NES | P-value | Tissue-FDR | Global-FDR |
| --- | --- | --- | --- | --- | --- | --- |
| Postsynaptic ribosome | Cerebellar hemisphere | 23 | -1.6 | 0.02 | 0.63 | 1.0 |
| Postsynaptic membrane | Frontal cortex | 15 | -1.6 | 0.02 | 0.36 | 1.0 |
| integral_component_of_presynaptic_membrane | Caudate basal ganglia | 18 | 1.6 | 0.02 | 0.50 | 1.0 |
| Postsynaptic ribosome | Cerebellum | 23 | -1.6 | 0.03 | 0.92 | 1.0 |
| Presynaptic membrane | Caudate basal ganglia | 24 | 1.5 | 0.05 | 0.52 | 1.0 |

Footnote: only top 5 pathways with smallest p-values were shown due to no significant

### Supplementary Table 14. Number of genes included in each brain tIssue in GTEx v8

| GTEx brain tissue | Number of genes included in Hong Kong cohort | Number of genes included in STRATA-G cohort |
| --- | --- | --- |
| Amygdala | 2667 | 2455 |
| Anterior cingulate cortex | 3385 | 3122 |
| Caudate basal ganglia | 4784 | 4441 |
| Cerebellar Hemisphere | 5529 | 5141 |
| Cerebellum | 6551 | 6056 |
| Cortex | 5275 | 4885 |
| Frontal cortex | 4365 | 4022 |
| Hippocampus | 3506 | 3186 |
| Hypothalamus | 1669 | 3180 |
| Nucleus accumbent basal ganglia | 4621 | 4257 |
| Putamen basal ganglia | 4223 | 3930 |
| Spinal cord cervical | 3118 | 2867 |
| Substantia nigra | 2439 | 2259 |

### Supplementary Table 15. Number of subjects in each individual cohort in STRATA-G

| Contributing site/study | TRS | Non-TRS |
| --- | --- | --- |
| AESOP London | 17 | 37 |
| Belfast | 13 | 83 |
| Bologna | 2 | 34 |
| GAP London | 30 | 125 |
| Istanbul | 17 | 26 |
| Lausanne | 18 | 180 |
| Oslo | 3 | 117 |
| Paris | 6 | 25 |
| Prague | 9 | 34 |
| Santander | 47 | 308 |
| Sao Paulo | 2 | 41 |
| UCL | 4 | 65 |
| Total | 168 | 1075 |

### Supplementary Table 16. Details of 107 synaptic pathways out of 259 from SynGo (geneset sizes between 15-500 unique genes) included in this study, listed in alphabetical order

| anchored component of synaptic vesicle membrane | neurotransmitter receptor localization to postsynaptic specialization membrane | postsynaptic ribosome | protein catabolic process at postsynapse | regulation of synaptic vesicle endocytosis | trans-synaptic signaling |
| --- | --- | --- | --- | --- | --- |
| axo-dendritic transport | neurotransmitter receptor transport | postsynaptic signaling pathway | protein catabolic process at synapse | regulation of synaptic vesicle exocytosis | translation at postsynapse |
| chemical synaptic transmission | neurotransmitter receptor transport to plasma membrane | postsynaptic specialization | regulation of modification of postsynaptic actin cytoskeleton | regulation protein catabolic process at postsynapse | translation at presynapse |
| dendritic transport | postsynapse organization | postsynaptic specialization assembly | regulation of modification of postsynaptic structure | structural constituent of postsynapse | translation at synapse |
| extrinsic component of synaptic vesicle membrane | postsynapse to nucleus signaling pathway | postsynaptic specialization membrane | regulation of neurotransmitter receptor localization to postsynaptic specialization membrane | structural constituent of synapse | transmitter-gated ion channel activity involved in regulation of postsynaptic membrane potential |
| integral component of postsynaptic density membrane | postsynaptic actin cytoskeleton | presynapse assembly | regulation of postsynapse assembly | synapse adhesion between pre- and post-synapse | Synaptic transport |
| integral component of postsynaptic membrane | postsynaptic actin cytoskeleton organization | presynaptic active zone | regulation of postsynapse organization | synapse assembly | voltage-gated ion channel activity involved in regulation of presynaptic membrane potential |
| integral component of postsynaptic specialization membrane | postsynaptic cytoskeleton | presynaptic active zone cytoplasmic component | regulation of postsynaptic cytosolic calcium levels | synapse maturation |  |
| integral component of presynaptic active zone membrane | postsynaptic cytoskeleton organization | presynaptic active zone membrane | regulation of postsynaptic density assembly | synapse organization |  |
| integral component of presynaptic membrane | postsynaptic cytosol | presynaptic cytosol | regulation of postsynaptic membrane neurotransmitter receptor levels | synaptic cleft |  |
| integral component of synaptic membrane | postsynaptic density assembly | presynaptic dense core vesicle exocytosis | regulation of postsynaptic membrane potential | synaptic membrane |  |
| integral component of synaptic vesicle membrane | postsynaptic density membrane | presynaptic endocytic zone | regulation of postsynaptic neurotransmitter receptor activity | synaptic signaling |  |
| ligand-gated ion channel activity involved in regulation of presynaptic membrane potential | postsynaptic density, intracellular component | presynaptic endocytic zone membrane | regulation of postsynaptic neurotransmitter receptor endocytosis | synaptic vesicle |  |
| maintenance of synapse structure | postsynaptic endocytic zone | presynaptic endosome | regulation of presynapse assembly | synaptic vesicle cycle |  |
| Synaptic metabolism | postsynaptic endosome | presynaptic membrane | regulation of presynaptic cytosolic calcium levels | synaptic vesicle endocytosis |  |
| modification of postsynaptic actin cytoskeleton | postsynaptic membrane | presynaptic modulation of chemical synaptic transmission | regulation of presynaptic membrane potential | synaptic vesicle endosomal processing |  |
| modification of postsynaptic structure | postsynaptic modulation of chemical synaptic transmission | presynaptic process involved in chemical synaptic transmission | regulation of synapse assembly | synaptic vesicle exocytosis |  |
| modification of synaptic structure | postsynaptic neurotransmitter receptor endocytosis | presynaptic ribosome | regulation of synapse maturation | synaptic vesicle membrane |  |
| modulation of chemical synaptic transmission | postsynaptic process involved in chemical synaptic transmission | process in the postsynapse | regulation of synapse organization | synaptic vesicle priming |  |
| neuronal dense core vesicle | postsynaptic recycling endosome | process in the presynapse | regulation of synaptic vesicle cycle | synaptic vesicle proton loading |  |

### Supplementary Table 17. Details of 127 additional brain-related pathways included from Go:BP (i.e. geneset sizes between 15-500 genes), by alphabetical order

| Keyword: Axon* | | | | |
| --- | --- | --- | --- | --- |
| GOBP anterograde axonal transport | GOBP axon ensheathment in central nervous system | GOBP axon extension | GOBP axonal fasciculation | GOBP axonal transport |
| GOBP axonal transport of mitochondrion | GOBP axonemal dynein complex assembly | GOBP axoneme assembly | GOBP central nervous system neuron axonogenesis | GOBP central nervous system projection neuron axonogenesis |
| GOBP motor neuron axon guidance | GOBP negative regulation of axon extension | GOBP negative regulation of axon extension involved in axon guidance | GOBP negative regulation of axonogenesis | GOBP positive regulation of axon extension |
| GOBP positive regulation of axonogenesis | GOBP regulation of axonogenesis | GOBP response to axon injury | GOBP retinal ganglion cell axon guidance | GOBP retrograde axonal transport |
| Keyword: Dendrite | | | | |
| GOBP dendrite development | GOBP dendrite extension | GOBP dendrite morphogenesis | GOBP dendrite self-avoidance | GOBP positive regulation of dendrite development |
| GOBP positive regulation of dendrite morphogenesis | GOBP regulation of dendrite development | GOBP regulation of dendrite extension | GOBP regulation of dendrite morphogenesis |  |
| Keyword: Glial | | | | |
| GOBP Glial cell apoptotic process | GOBP glial cell development | GOBP glial cell differentiation | GOBP glial cell migration | GOBP glial cell proliferation |
| GOBP microglial cell activation | GOBP negative regulation of glial cell differentiation | GOBP positive regulation of glial cell differentiation | GOBP positive regulation of glial cell proliferation | GOBP regulation of glial cell differentiation |
| GOBP regulation of glial cell migration | GOBP regulation of glial cell proliferation | GOBP regulation of microglial cell activation | GOBP telencephalon glial cell migration |  |
| Keyword: Neuron* | | | | |
| GOBP central nervous system neuron development | GOBP central nervous system neuron differentiation | GOBP cerebral cortex neuron differentiation | GOBP dopaminergic neuron differentiation | GOBP ensheathment of neurons |
| GOBP forebrain generation of neurons | GOBP forebrain neuron development | GOBP forebrain neuron differentiation | GOBP midbrain dopaminergic neuron differentiation | GOBP negative regulation of neuron apoptotic process |
| GOBP negative regulation of neuron death | GOBP negative regulation of neuron differentiation | GOBP negative regulation of neuron projection development | GOBP negative regulation of neuron regeneration | GOBP negative regulation of oxidative stress induced neuron death |
| GOBP neuron apoptotic process | GOBP neuron cell cell adhesion | GOBP neuron cellular homeostasis | GOBP neuron death | GOBP neuron death in response to oxidative stress |
| GOBP neuron fate commitment | GOBP neuron fate specification | GOBP neuron maturation | GOBP neuron migration | GOBP neuron projection arborization |
| GOBP neuron projection extension | GOBP neuron projection extension involved in neuron projection | GOBP neuron projection guidance | GOBP neuron projection organization | GOBP neuron projection regeneration |
| GOBP neuron recognition | GOBP neuronal action potential | GOBP neuronal stem cell population maintenance | GOBP positive regulation of neuron apoptotic process | GOBP positive regulation of neuron death |
| GOBP positive regulation of neuron differentiation | GOBP positive regulation of neuron migration | GOBP positive regulation of neuron projection development | GOBP regulation of long term neuronal synaptic plasticity | GOBP regulation of neuron differentiation |
| GOBP regulation of neuron migration | GOBP regulation of projection development | GOBP regulation of projection regeneration | GOBP regulation of neuronal synaptic plasticity | GOBP semaphoring plexin signaling pathway involved in neuron projection guidance |
| GOBP spinal cord motor neuron differentiation | GOBP wnt signaling pathway involved in midbrain dopaminergic neuron differentiation |  |  |  |

### Supplementary Figure 1. An undirected network graph showing the pairwise similarity between pairs of pathways using Jaccard index.

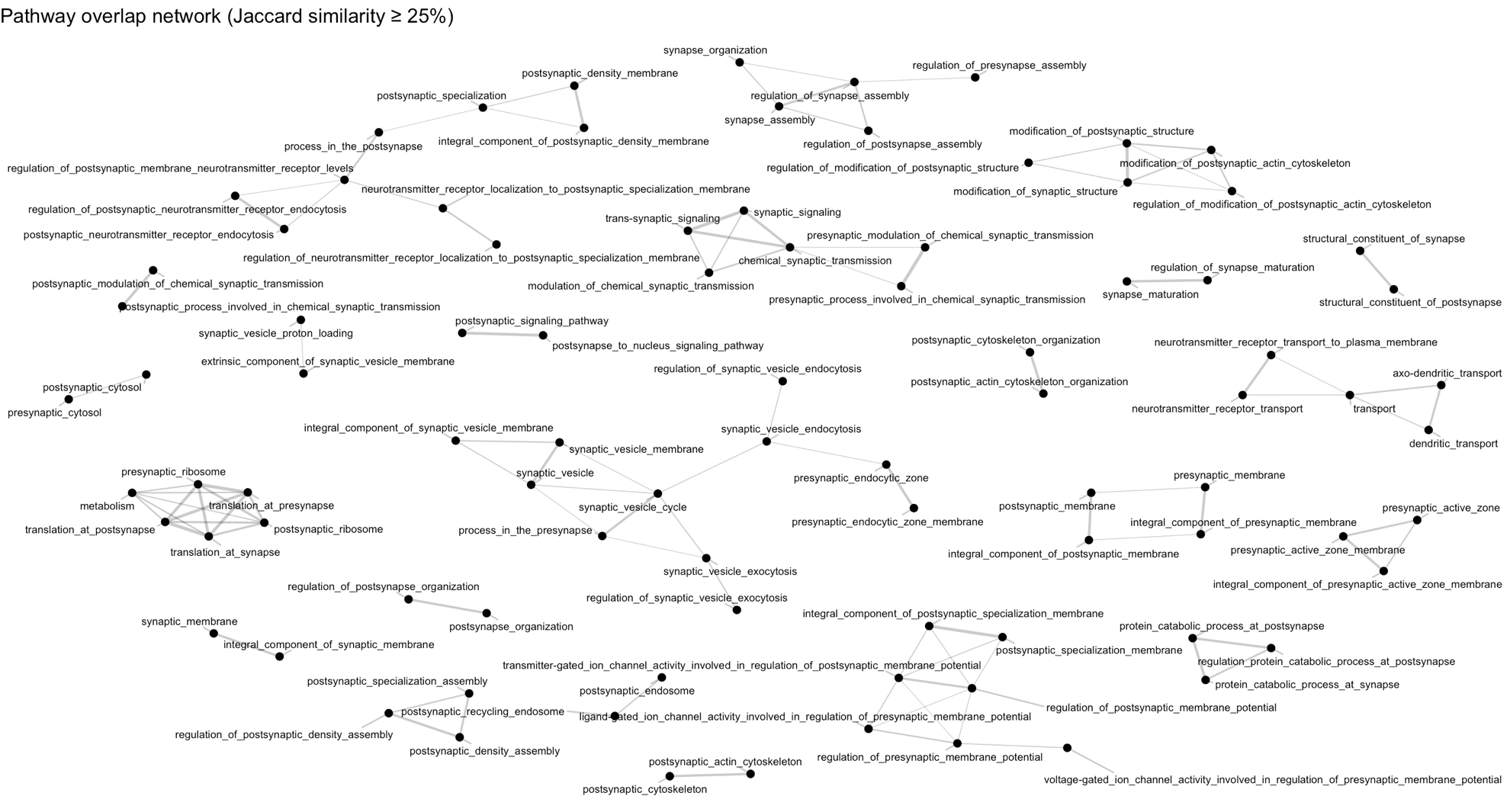

### Supplementary Figure 2. Variant calling, imputation, and QC steps in Hong Kong cohort

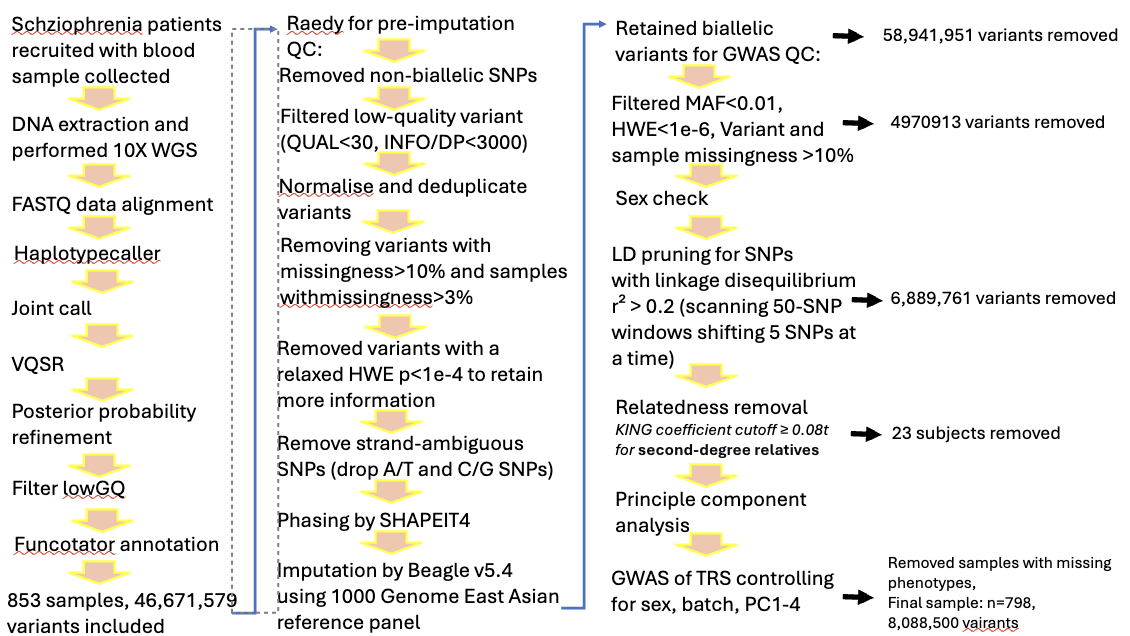

### Supplementary Figure 3. Manhattan plot of GWAS of Hong Kong sample

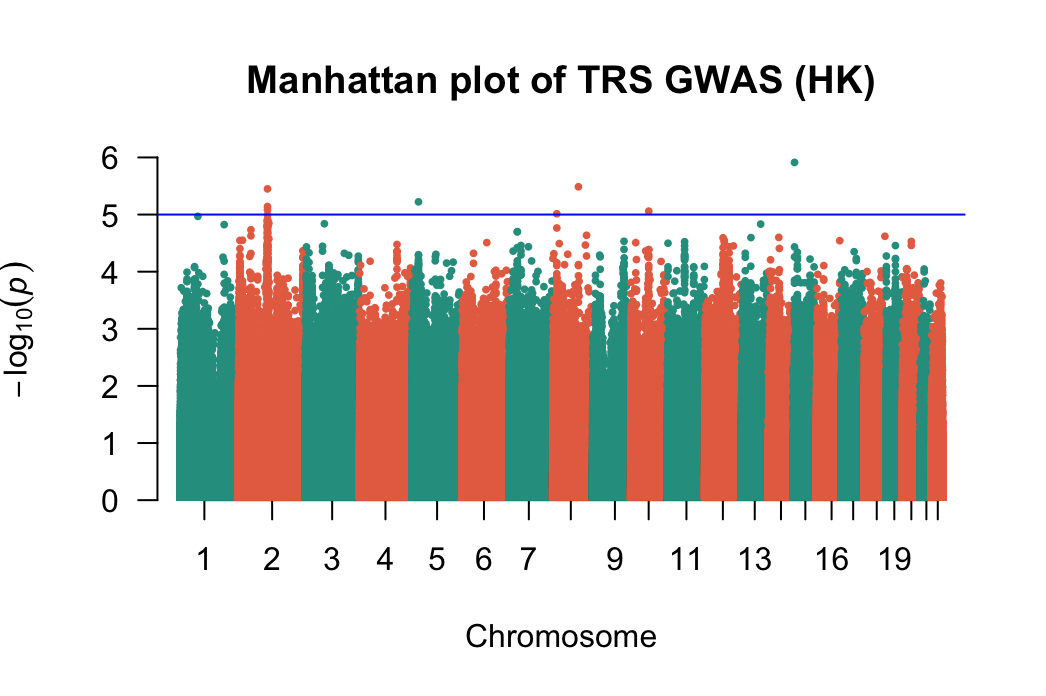

### Supplementary Figure 4. QQ plot of GWAS of Hong Kong sample

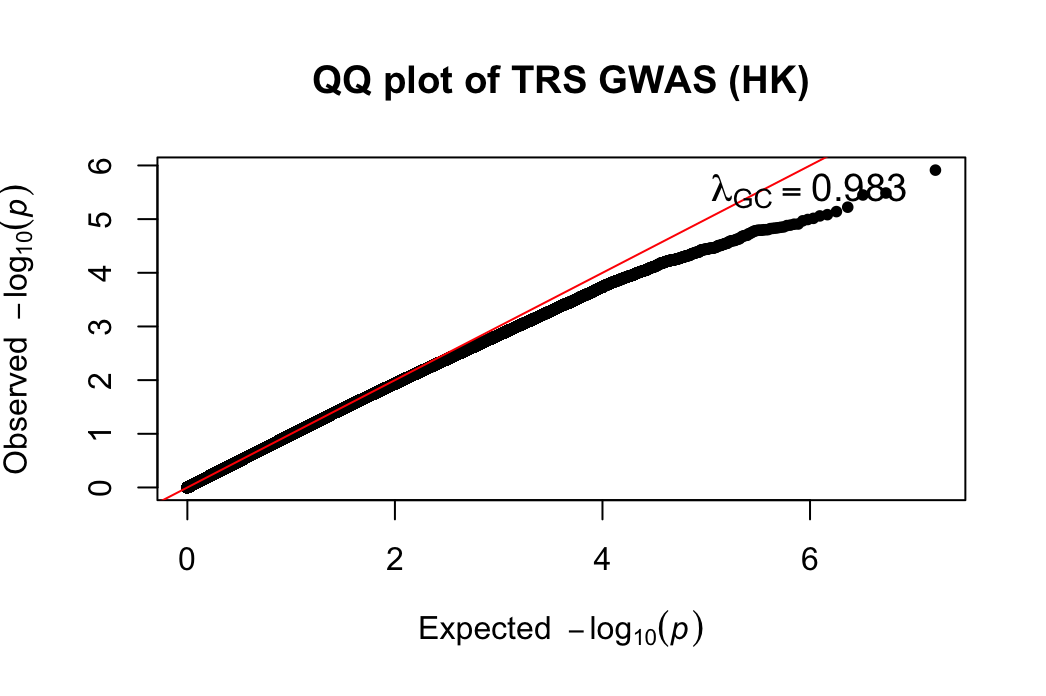

### Supplementary Figure 5. Manhattan plot of GWAS of STRATA-G sample

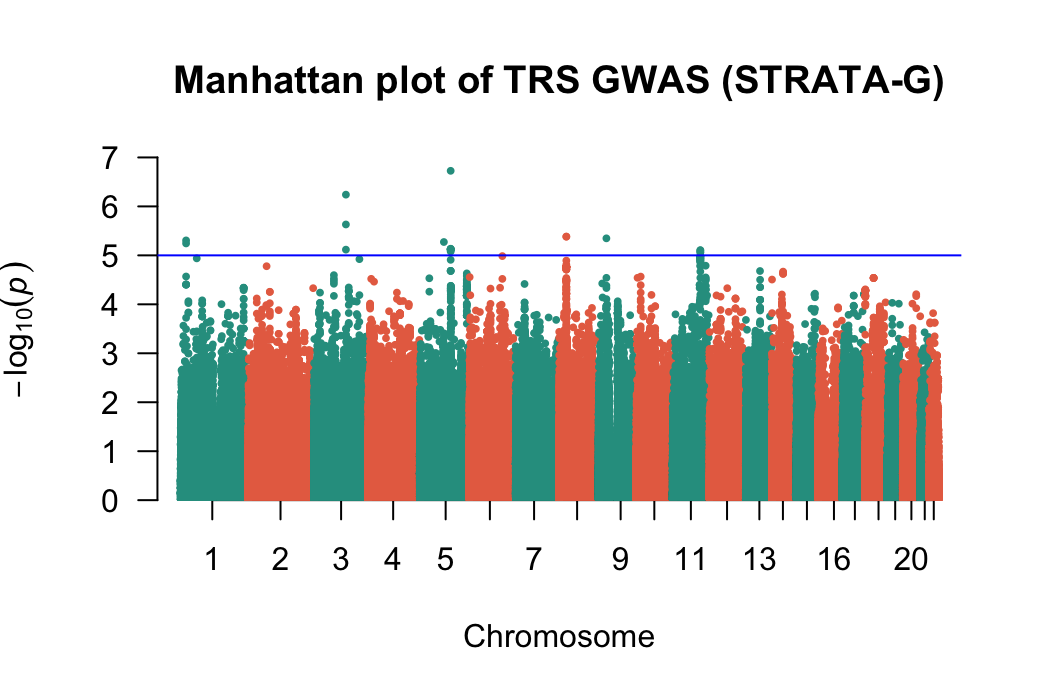

### Supplementary Figure 6. QQ plot of GWAS of STRATA-G sample

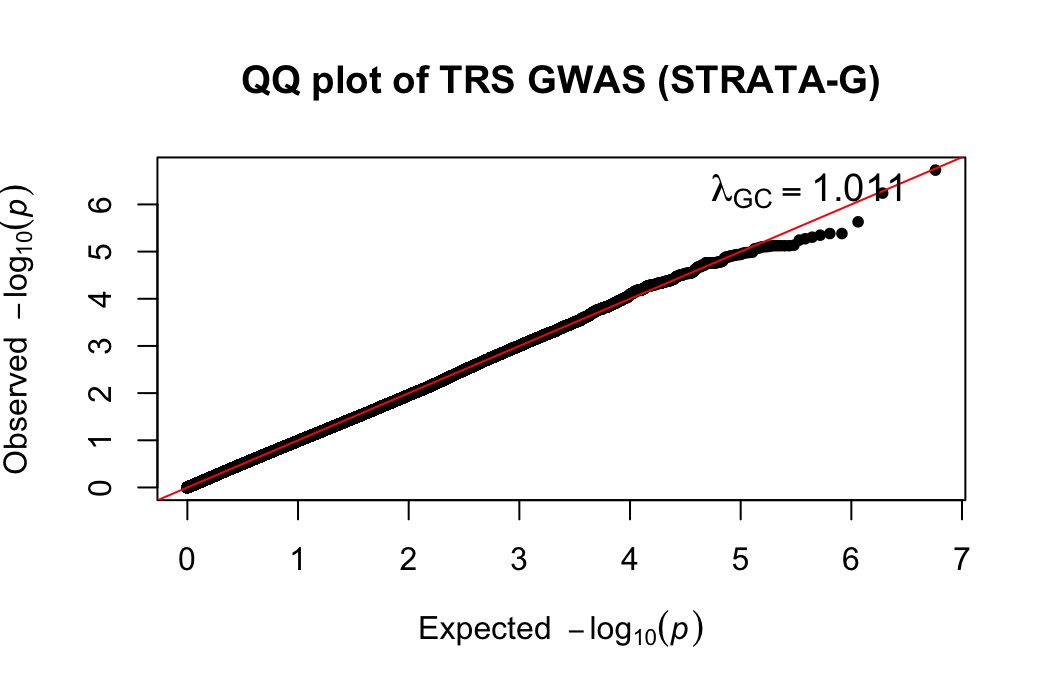

### Supplementary Figure 7. Manhattan plot of meta-analysed GWAS (Hong Kong + STRATA-G)

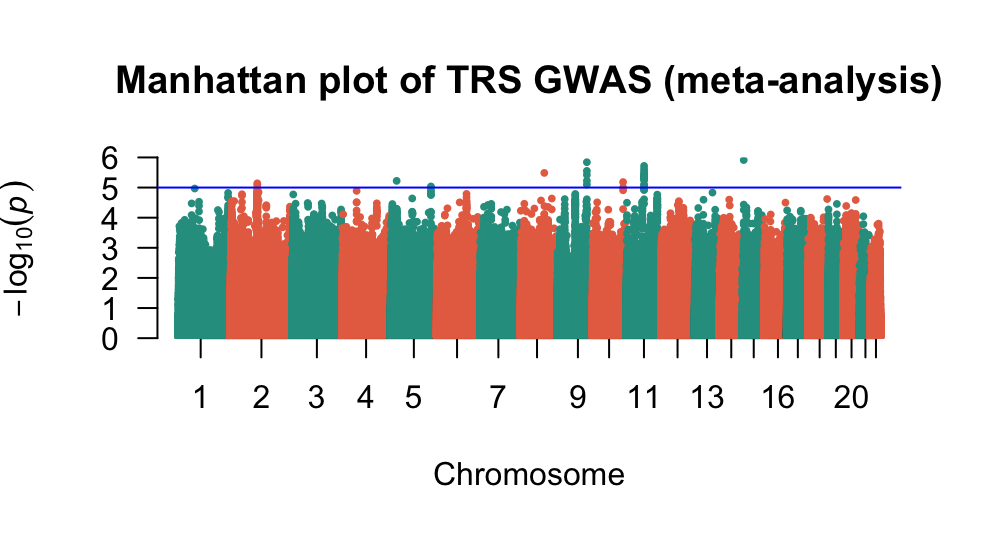

### Supplementary Figure 8. QQ plot of meta-analysed GWAS (Hong Kong + STRATA-G)

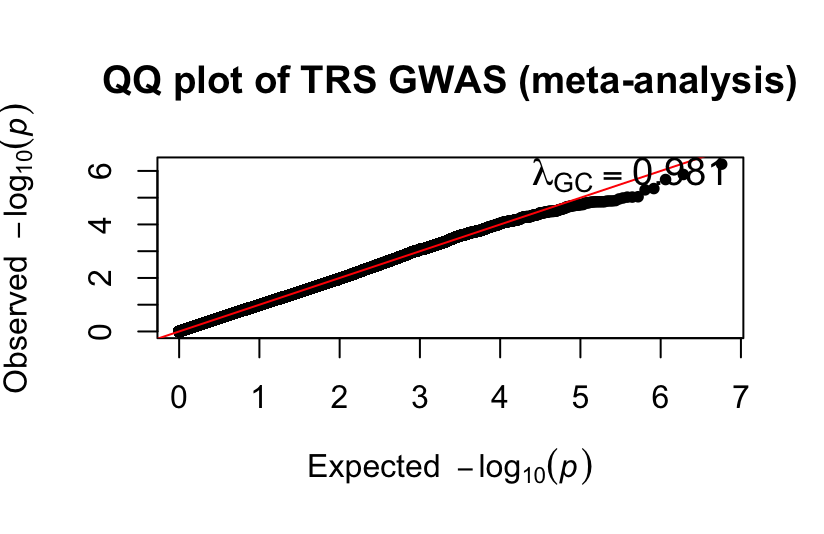

### Supplementary Figure 9. Quintile plots of SCZ-PRS (PRSCSeur-hk, PRSCSeas-hk, PRSCSx-hk, PRSCSeur-strata-G)

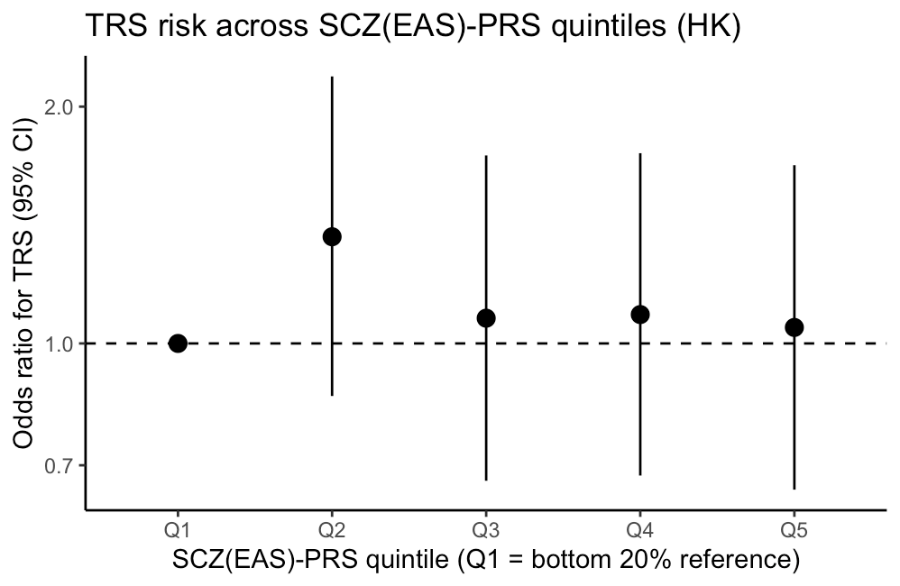

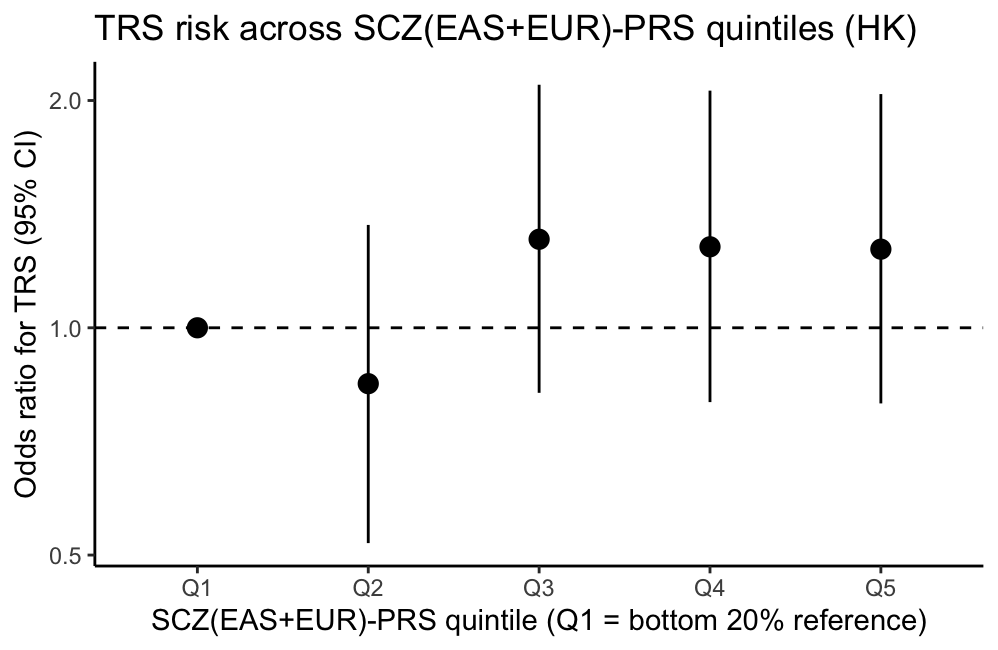

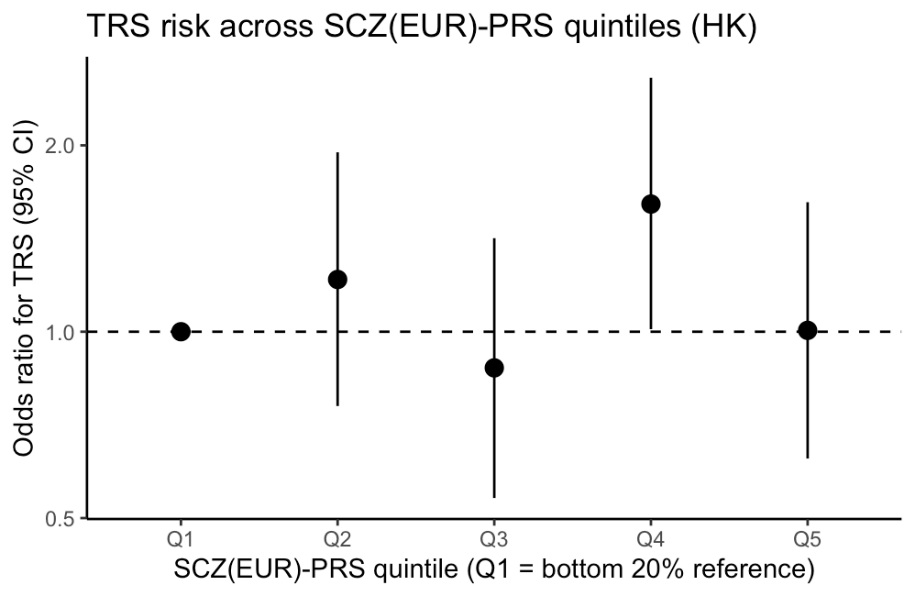

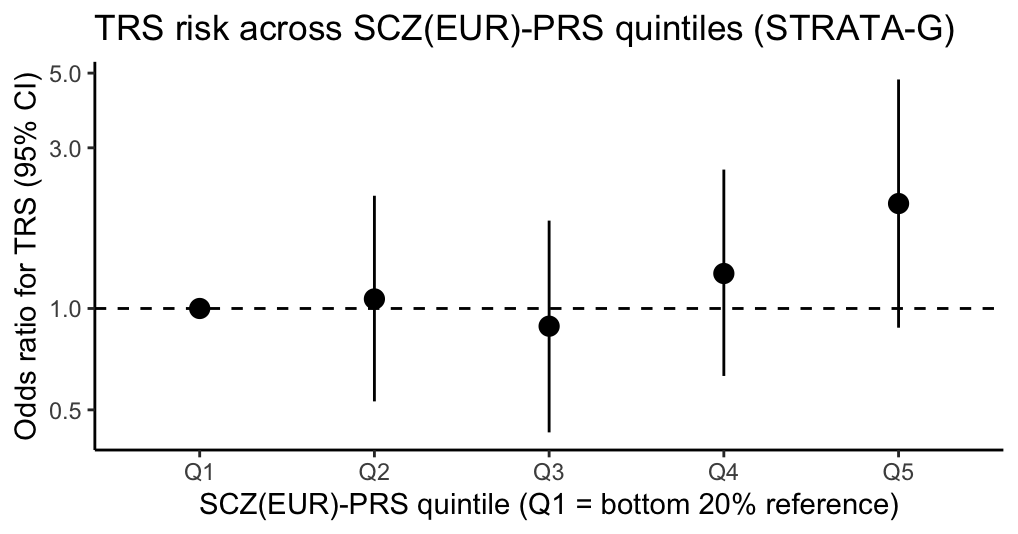

### Supplementary Figure 10. Violin plot of SCZ-PRS (PRSCSeur-hk, PRSCSeas-hk, PRSCSx-hk, PRSCSeur-strata-G)

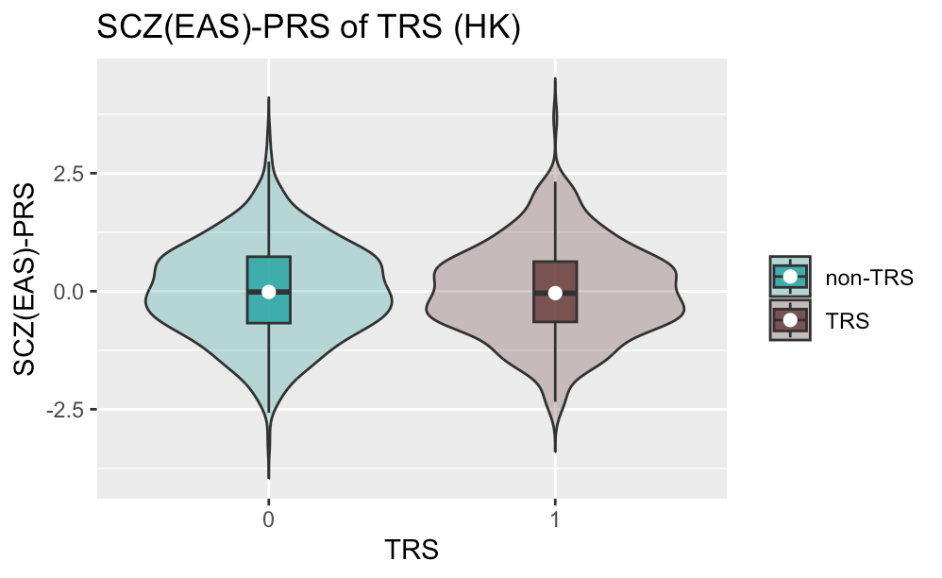

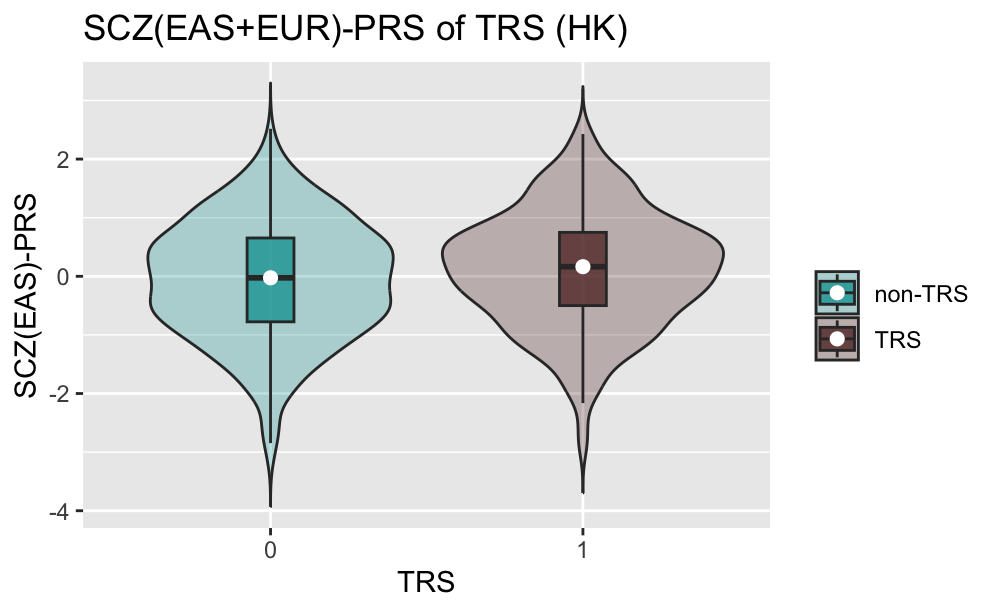

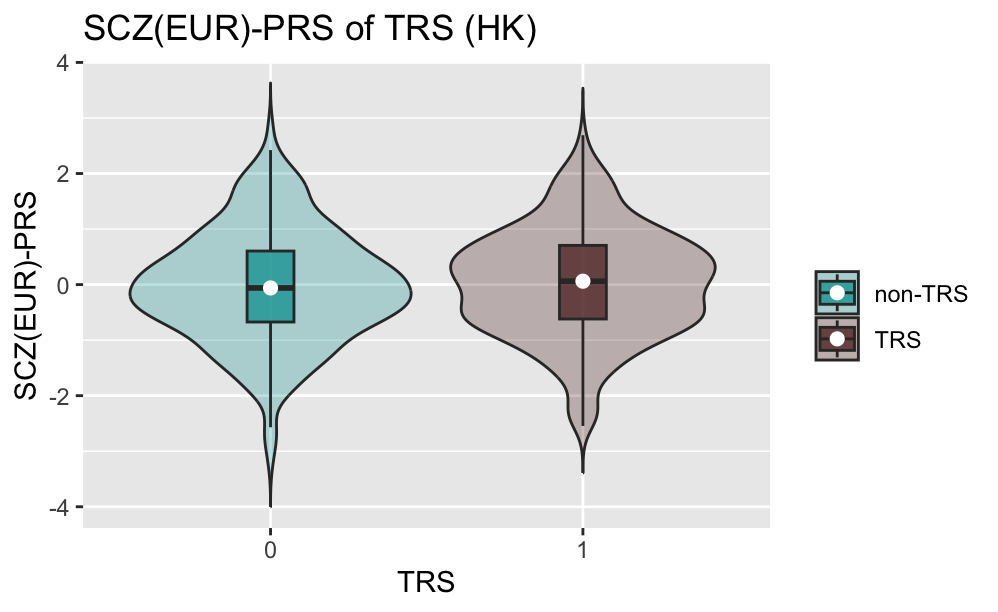

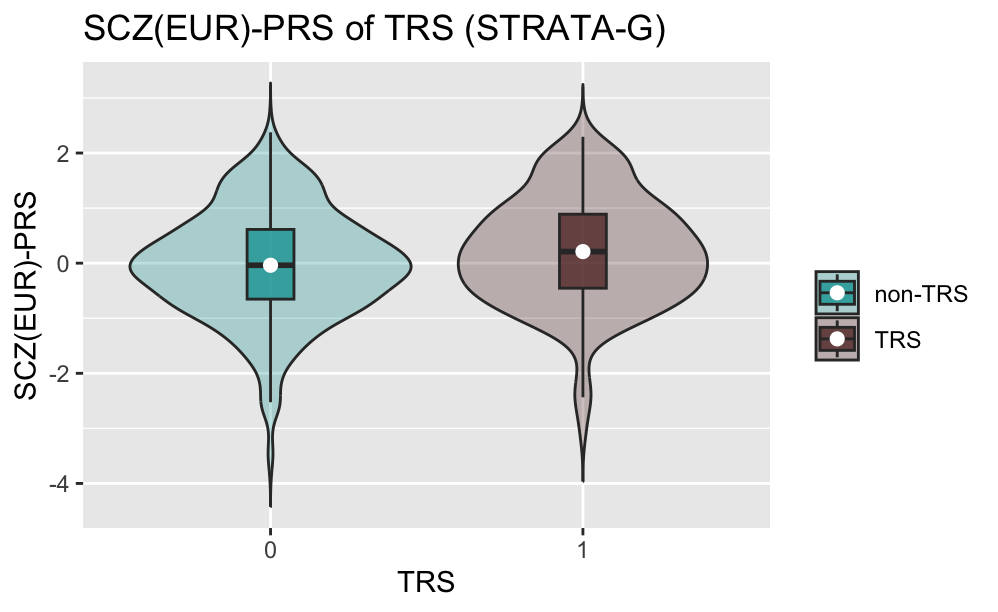

### Supplementary Figure 11. Quintile plot of TRS-PRS (PRSCS-hk, PRSCS-strataG)

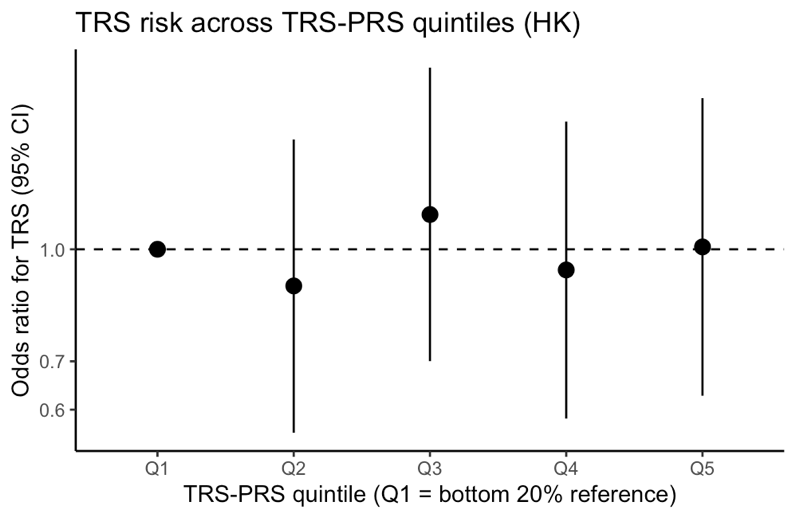

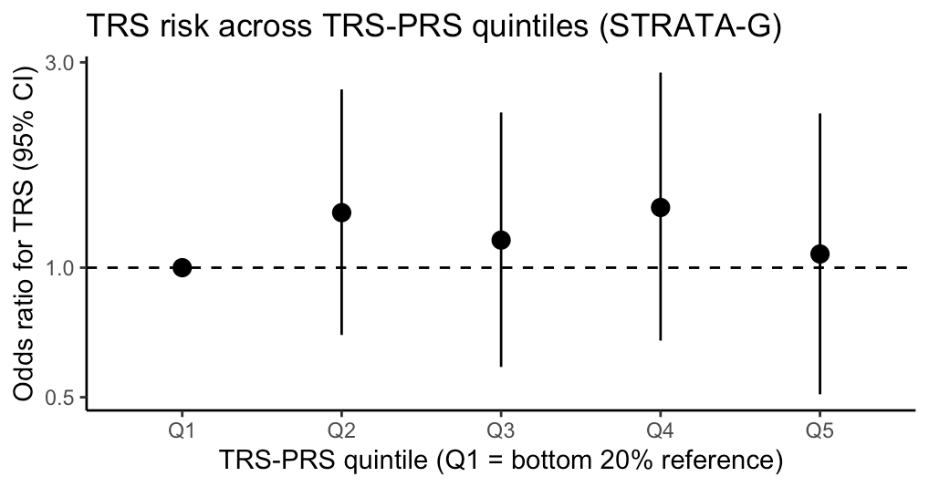

### Supplementary Figure 12. Violin plot of TRS-PRS (PRSCS-hk, PRSCS-strataG)

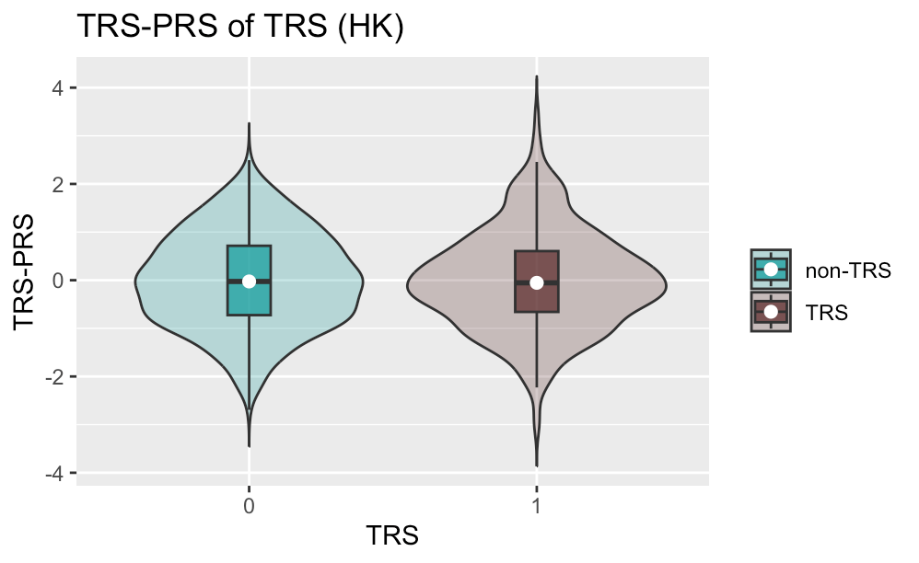

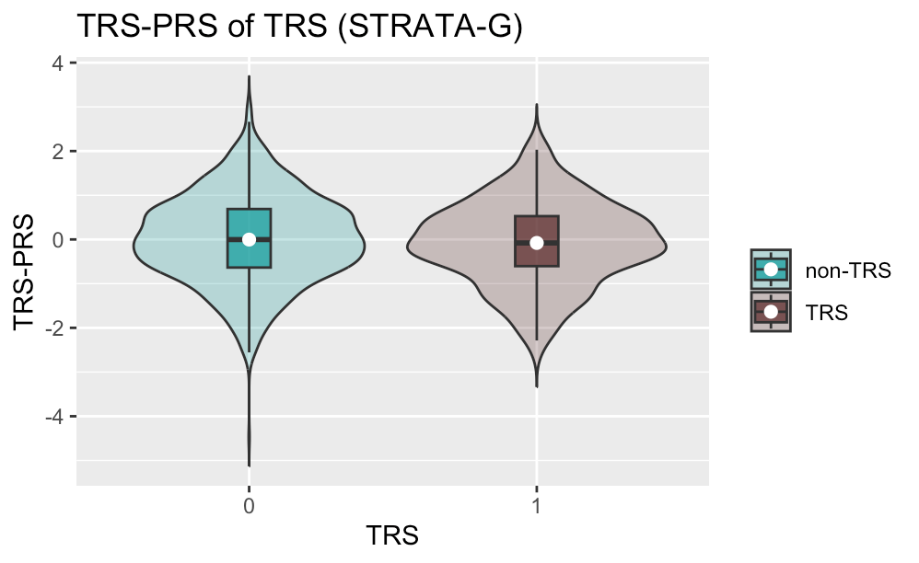

### Supplementary Figure 13. Violin plot of pPRS

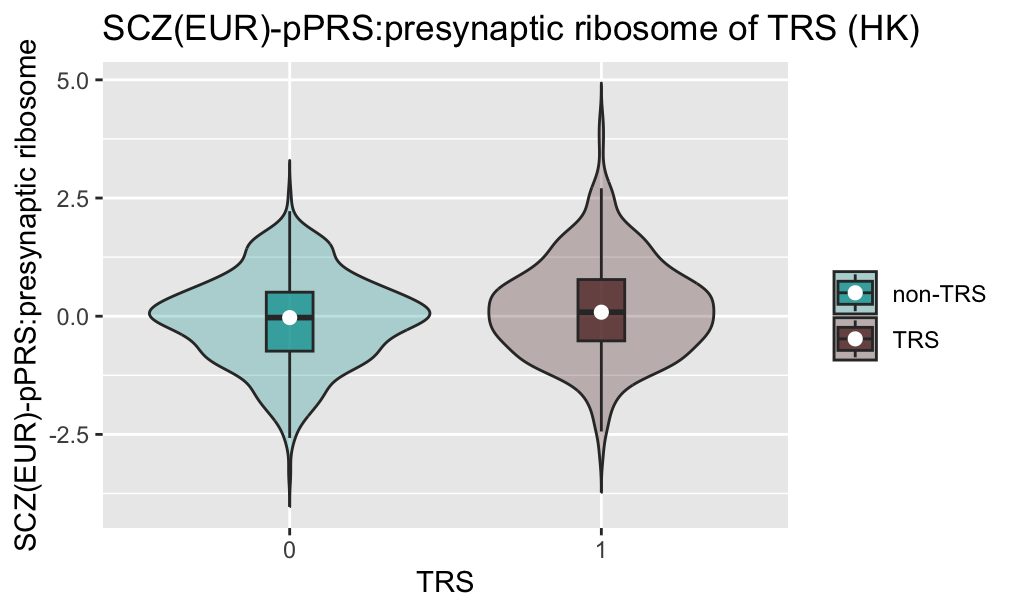

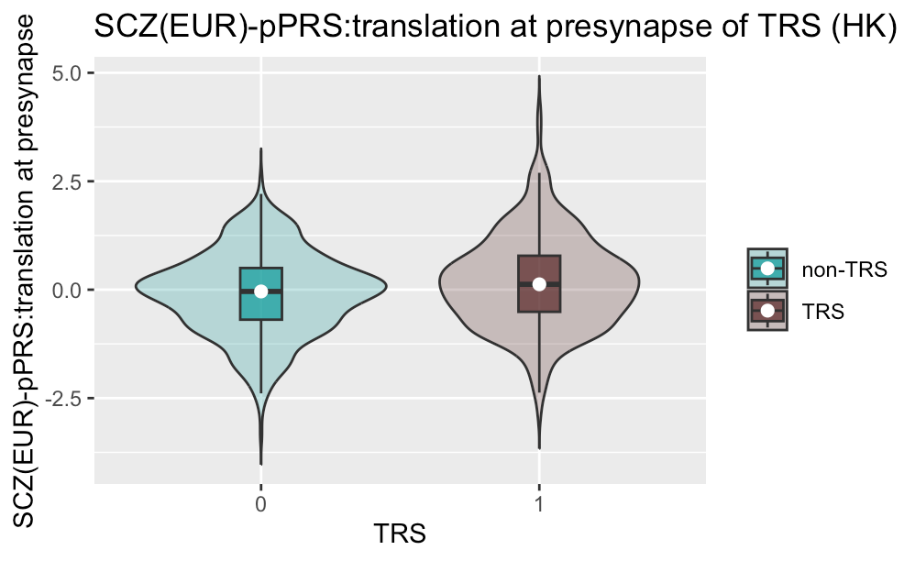

### Supplementary Figure 14. Quintile plot of pPRS

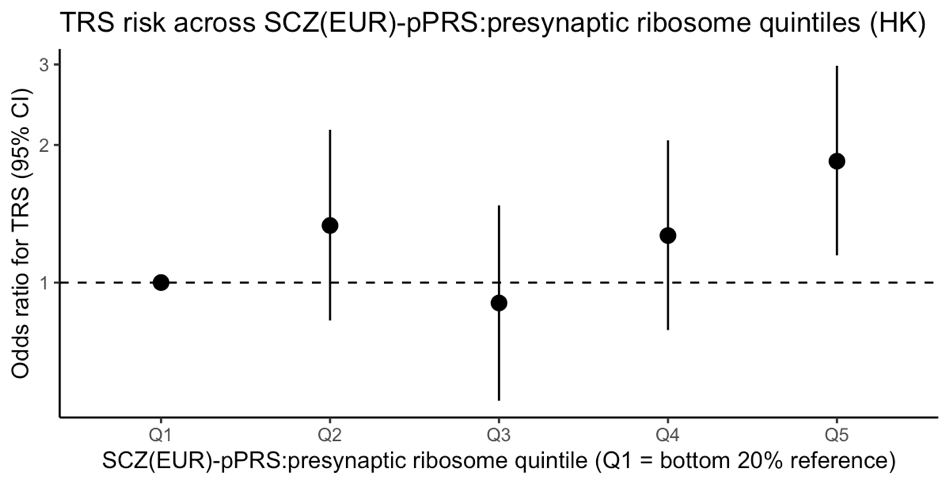

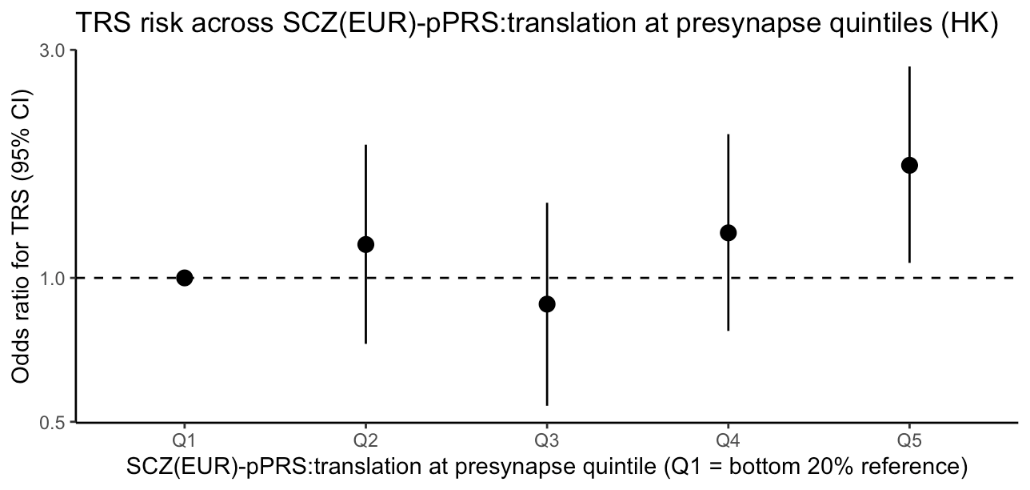

### Supplementary Figure 15. Elbow plot of PCA in Hong Kong sample

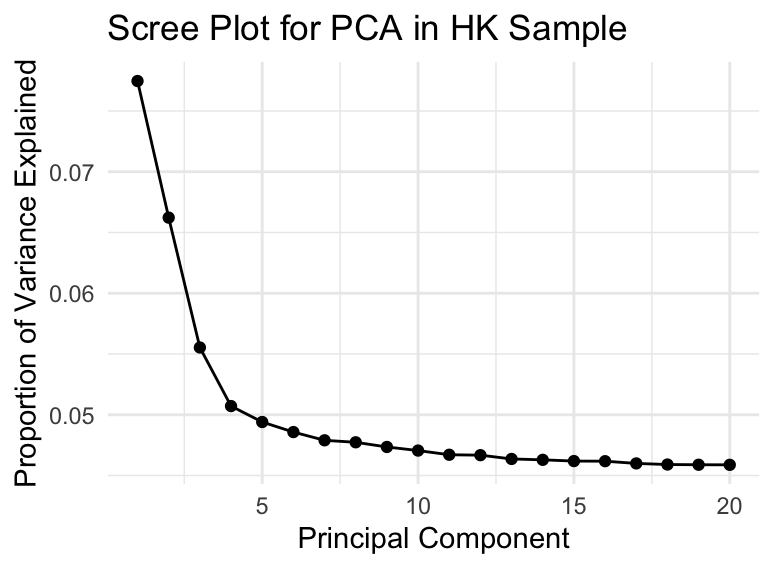

### Supplementary Figure 16. Elbow plot of PCA in STRATA-G sample

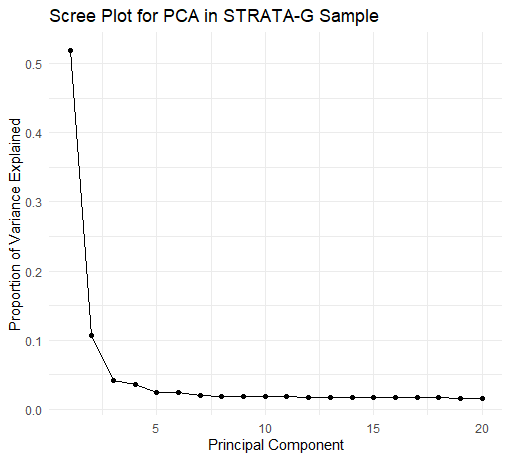

### Supplementary Figure 17. PC1 vs PC2 and PC1 vs PC3 panels in both Hong Kong and STRATA-G sample

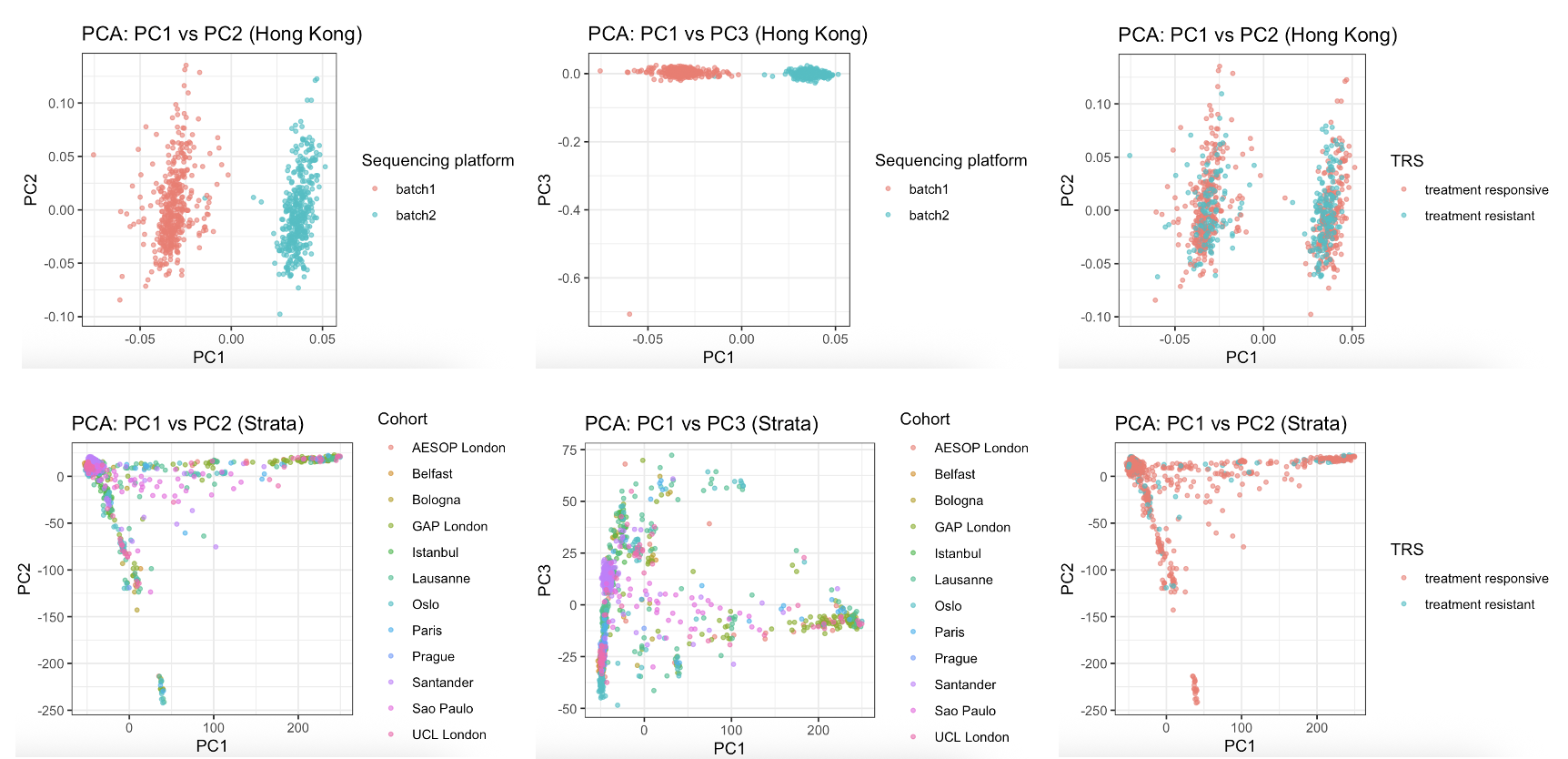
Footnote: In HK cohort, PC1 primarily reflects sequencing platform, while higher-order PCs capture residual population structure given no ancestrial differences.

### Supplementary Method. DNA collection, extraction, and sequencing method

DNA was extracted from EDTA-venous blood samples using standard extraction procedure involving cell lysis and purification. DNA quantity and quality were assessed using spectrophotometric and fluorometric methods. Samples passing quality control (OD ratio: 260/280, >= 1.7]) were used for library preparation and whole-genome sequencing. Sequencing was performed to a mean depth of approximately 10x in three batches using MGI DNBSEQ-T7RS sequencing platforms following standard protocols.
